# Proteomic analysis of hepatocellular carcinoma etiology and risk stratification in two prospective studies

**DOI:** 10.64898/2025.12.08.25341832

**Authors:** Eleanor L. Watts, Cody Z. Watling, Li C. Cheung, Christian C. Abnet, Belynda Hicks, Wen-Yi Huang, Amy A. Hutchinson, Hormuzd Katki, Erikka Loftfield, Mitchell J. Machiela, Katherine A. McGlynn, Jianxin Shi, Rachael Stolzenberg-Solomon, Steven C. Moore

## Abstract

High-throughput proteomics enable deeper insights into hepatocellular carcinoma (HCC) etiology and risk stratification. In a nested study within Prostate, Lung, Colorectal and Ovarian Screening trial (118 HCC cases, 118 controls; median follow-up=9.7 years), we examined 4,003 circulating proteins using conditional logistic regression and developed an eight-protein risk score via LASSO regression. Findings were validated in UK Biobank (50,182 participants; 36 HCC cases; median follow-up=13.9 years) using Cox regression and the C-index. In PLCO, 106 proteins were significantly associated with HCC risk; 97% of those available were replicated in UK Biobank, implicating inflammation and growth-factor signaling. The risk score demonstrated strong discrimination (C-index=0.92, 95% confidence intervals [CI] 0.84-0.97) overall and ability to stratify 8-year cumulative HCC risk in at-risk subgroups including cirrhosis (18% for higher risk vs 2% lower risk) or current/past hepatitis B/C infection (13% vs 1%). These findings support proteomic profiling for identifying the highest-risk individuals for precision surveillance.

## 1. Introduction

Liver cancer is the sixth most common cancer and the third-leading cause of cancer mortality worldwide^1^. Hepatocellular carcinoma (HCC), which accounts for 80% of liver cancers^2^, typically develops in the context of cirrhosis, driven by risk factors such as chronic hepatitis B virus (HBV) and hepatitis C virus (HCV) infections, heavy alcohol consumption, and increasingly, obesity, metabolic syndrome, and metabolic-associated steatotic liver disease (MASLD)^3^. However, the population attributable fraction accounts for approximately 60% of cases, suggesting that additional etiological factors remain unidentified^4^.

Prognosis for HCC is poor, with a 5-year survival of 20-30% in the US^5,6^. However, survival can reach 60-70% if diagnosed at an early stage, when treatment is most effective^7,8^. Surveillance is recommended only for individuals at highest risk, namely those with cirrhosis and/or chronic HBV infection, using bi-annual abdominal ultrasound, with or without alpha fetoprotein (AFP) testing^9,10^. However, this approach has low sensitivity for detecting early-stage HCC (63% for AFP with ultrasound), performs less well among individuals with obesity (a major cirrhosis risk factor)^11–13^, has poor adherence (<30% following recommended surveillance)^14^, and fails to account for individual differences in risk.

Recent proteomic studies have identified circulating proteins associated with HCC and liver cancer risk years before diagnosis, demonstrating their potential to clarify disease etiology^15,16^. Although many proteins show stability and predictive value across diseases, including HCC, most remain uninvestigated^16–21^. Moreover, few studies have assessed how long before diagnosis such predictive associations remain robust. To address these gaps, we analysed associations of over 4,000 circulating proteins with HCC risk in a nested case-control study within the Prostate, Lung, Colorectal and Ovarian (PLCO) Cancer Screening Trial, with independent validation in the UK Biobank.

## 2. Results

In PLCO, 118 HCC cases were matched to 118 controls, with a median time to diagnosis of 9.7 years. In the UK Biobank, 50,182 cancer-free participants were included in the proteomics subset, among whom 36 subsequently developed HCC, with a median time-to-diagnosis of 6.9 years. Participants who developed HCC had higher proportion of obesity (30+ kg/m^2^), diabetes, smoking and heavier alcohol consumption in both cohorts (**Table 1**). They also had a higher proportion of liver conditions. In PLCO, 7 cases (5.9%) reported a history of cirrhosis, and 17 (14.4%) reported hepatitis (any type), whereas no controls reported these conditions. Similarly, in UK Biobank, 17% of incident HCC cases had a history of cirrhosis, and 14% had evidence of current or past HBV or HCV infection at baseline, based on self-report and hospitalization records (**Table 1**).

**Table 1:** Participant characteristics in the PLCO and UK Biobank study populations.

|  | PLCO |  | UK Biobank |  |
| --- | --- | --- | --- | --- |
|  | Controls<br>(N=118) | HCC cases<br>(N=118) | Whole cohort<br>(N= 50,182) | HCC cases<br>(N=36) |
| Age at blood draw, yrs med (IQR) | 63.1 (8.6) | 63.2 (7.8) | 58.4 (13.4) | 61.9 (7.1) |
| Time to diagnosis, yrs med (IQR) | - | 9.7 (9.9) | - | 6.9 (6.4) |
| Sex |  |  |  |  |
| Men | 96 (81.4%) | 96 (81.4%) | 23,393 (46.6%) | 25 (69.4%) |
| Women | 22 (18.6%) | 22 (18.6%) | 26,789 (53.4%) | 11 (30.6%) |
| Race and ethnicity* |  |  |  |  |
| Asian/Pacific Islander | 9 (7.6%) | 9 (7.6%) | 1,111 (2.2%) | 1 (2.8%) |
| Black | 5 (4.2%) | 5 (4.2%) | 1,169 (2.3%) | 0 (0.0%) |
| Hispanic | 9 (7.6%) | 9 (7.6%) | NA | NA |
| White, non-Hispanic | 95 (80.5%) | 95 (80.5%) | 46,724 (93.1%) | 34 (94.4%) |
| Other/missing | 0 (0.0%) | 0 (0.0%) | 1,178 (2.3%) | 1 (2.8%) |
| Education |  |  |  |  |
| Up to high school | 33 (28%) | 35 (29.7%) | 10,872 (21.7%) | 5 (13.9%) |
| Post high school | 84 (71.2%) | 82 (69.5%) | 29,760 (59.3%) | 18 (50%) |
| Missing | 1 (0.8%) | 1 (0.8%) | 9,550 (19%) | 13 (36.1%) |
| Body mass index, kg/m<sup>2</sup> |  |  |  |  |
| <25 | 36 (30.5%) | 26 (22%) | 16,207 (32.3%) | 8 (22.2%) |
| 25-<30 | 57 (48.3%) | 50 (42.4%) | 21,484 (42.8%) | 10 (27.8%) |
| 30+ | 21 (17.8%) | 40 (33.9%) | 12,247 (24.4%) | 17 (47.2%) |
| Missing | 4 (3.4%) | 2 (1.7%) | 244 (0.5%) | 1 (2.8%) |
| Smoking status |  |  |  |  |
| Never | 39 (33.1%) | 26 (22%) | 27,230 (54.3%) | 11 (30.6%) |
| Former | 68 (57.6%) | 78 (66.1%) | 17,367 (34.6%) | 17 (47.2%) |
| Current | 11 (9.3%) | 14 (11.9%) | 5,345 (10.7%) | 8 (22.2%) |
| Missing | 0 (0.0%) | 0 (0.0%) | 240 (0.5%) | 0 (0.0%) |
| Current alcohol consumption, drink/d† |  |  |  |  |
| None | 19 (16.1%) | 16 (13.6%) | 4,338 (8.6%) | 3 (8.3%) |
| ≤1 | 64 (54.2%) | 53 (44.9%) | 26,566 (52.9%) | 16 (44.4%) |
| >1-2 | 19 (16.1%) | 13 (11%) | 10,688 (21.3%) | 7 (19.4%) |
| >2 | 14 (11.9%) | 20 (16.9%) | 8,471 (16.9%) | 10 (27.8%) |
| Missing | 2 (1.7%) | 16 (13.6%) | 119 (0.2%) | 0 (0.0%) |
| Coffee consumption (cups/d) |  |  |  |  |
| None | 11 (9.3%) | 7 (5.9%) | 11,260 (22.4%) | 7 (19.4%) |
| >0-2 | 62 (52.5%) | 50 (42.4%) | 23,079 (46%) | 23 (63.9%) |
| >2 | 43 (36.4%) | 45 (38.1%) | 15,642 (31.2%) | 6 (16.7%) |
| Missing | 2 (1.7%) | 16 (13.6%) | 201 (0.4%) | 0 (0.0%) |
| Diabetes (self-report) |  |  |  |  |
| No | 107 (90.7%) | 85 (72%) | 47,133 (93.9%) | 26 (72.2%) |
| Yes | 11 (9.3%) | 32 (27.1%) | 2,828 (5.6%) | 10 (27.8%) |
| Missing | 0 (0.0%) | 1 (0.8%) | 221 (0.4%) | 0 (0.0%) |
| Liver cirrhosis at baseline‡ |  |  |  |  |
| No | 118 (100%) | 109 (92.4%) | 50,091 (99.8%) | 30 (83.3%) |
| Yes | 0 (0.0%) | 7 (5.9%) | 91 (0.2%) | 6 (16.7%) |
| Missing | 0 (0.0%) | 2 (1.7%) | 0 (0.0%) | 0 (0.0%) |
| Hepatitis at baseline‡ |  |  |  |  |
| No | 118 (100%) | 100 (84.7%) | 50,023 (99.7%) | 31 (86.1%) |
| Yes | 0 (0.0%) | 17 (14.4%) | 159 (0.3%) | 5 (13.9%) |
| Missing | 0 (0.0%) | 1 (0.8%) | 0 (0.0%) | 0 (0.0%) |

Values are n (%), unless otherwise indicated. PLCO controls were matched to cases based on age at blood draw, sex, race/ethnicity, study year of blood draw, and year of randomization.
\*Racial and ethnic groups are based on PLCO classifications. Corresponding UK Biobank categories: Asian/Pacific Islander → Asian or Asian British; Black → Black or Black British; Hispanic → not available; non-Hispanic White → White; Other/Missing → Mixed, Other, or White.
†Standard drink = 14 g pure alcohol (U.S. definition); UK Biobank values standardized to this metric.
‡PLCO: self-report, UK Biobank, self-report or hospital admission prior to study baseline
Abbreviations: HBV=hepatitis B virus, HCC=hepatocellular carcinoma, HCV=hepatitis C virus, IQR=interquartile range, PLCO=Prostate, Lung, Colorectal and Ovarian Cancer Screening Trial.

### 2.1. Protein associations with hepatocellular carcinoma

In PLCO, 106 proteins were significantly associated with HCC risk after Bonferroni correction (p<1.25×10^-5^); 105 showed positive associations. The strongest positive associations, ranked by significance, were tumor necrosis factor receptor superfamily member 11B (TNFRSF11B: OR per 1 SD=2.86, 95% CI 1.91-4.30), CUB domain-containing protein 1 (CDCP1: 3.76, 2.25-6.28), and carboxypeptidase Z (CPZ: 2.76, 1.85-4.11). Insulin-like growth factor-binding protein 3 (IGFBP-3) was the only protein inversely associated (0.35, 0.23-0.54; **Figure 1A**). Of these, 99 proteins were available in UK Biobank, and 96 (97%) exhibited directionally consistent associations at p<0.05 (**Supplementary Table 1**). In UK Biobank, 342 proteins were significantly associated with HCC after multiple testing correction (p<1.71×10^-5^, **Figure 1B**), including 82 that were also Bonferroni significant in PLCO (**Supplementary Data**).

**Figure 1:**
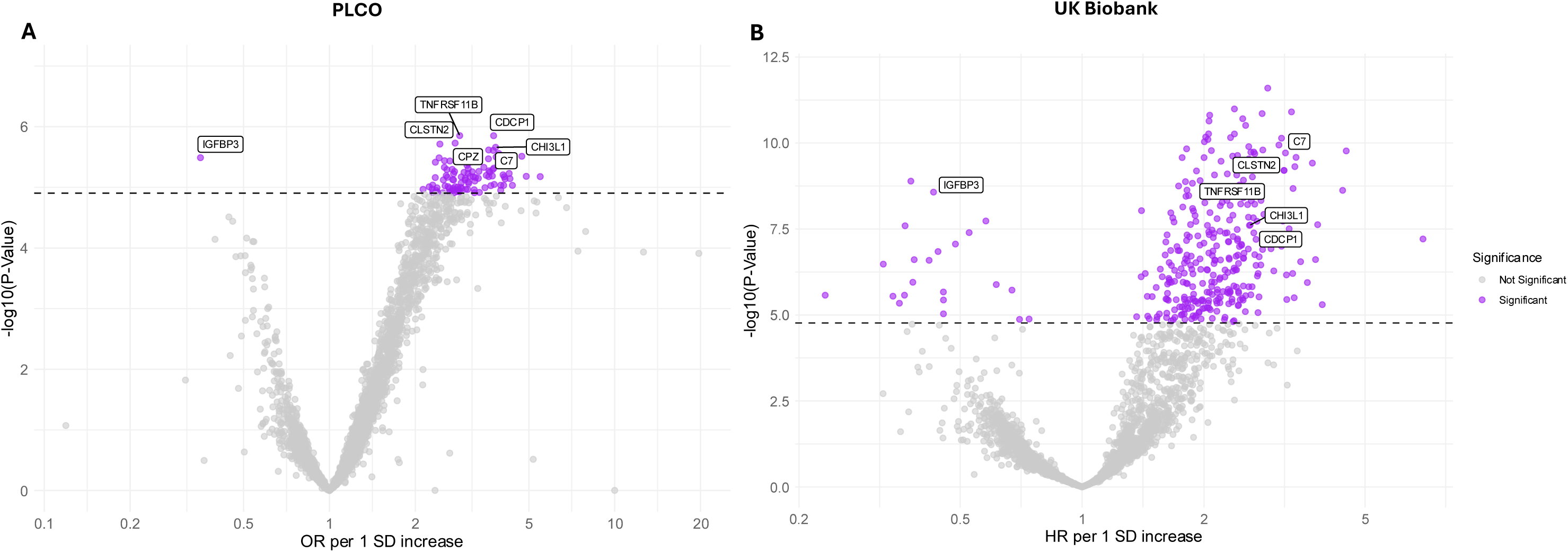
Volcano plots showing the associations of proteins per 1 SD increase with hepatocellular carcinoma risk. **A** PLCO: Conditional logistic regression models of each protein’s association with HCC, conditioned on matching factors, and adjusted for age, smoking, alcohol consumption, coffee consumption, education, BMI, diabetes. Missing covariates were imputed using MICE. Associations were corrected for multiple testing using Bonferroni (N tests=4,003, p<1.25×10^-5^). **B** UK Biobank: Cox regression models of each protein’s association with HCC, with follow-up time as the underlying variable. Models adjusted for age, sex, race, alcohol consumption, coffee consumption, smoking, education, BMI, and diabetes. Missing covariates were imputed using MICE. Associations were corrected for multiple testing using Bonferroni (N tests= 2,921, p<1.71×10^-5^). Labelled proteins are the top proteins ranked by lowest p-value in PLCO, CPZ was unavailable in UK Biobank. Associations for each protein and cohort are available from **Supplementary Data**. Abbreviations: ADGRE1=Adhesion G protein-coupled receptor E1, CDCP1=CUB domain-containing protein 1, CLSTN2=calsyntenin-2, CPZ=carboxypeptidase Z, HCC=hepatocellular carcinoma, HR=hazard ratio, IGFBP-3=insulin-like growth factor-binding protein 3, MICE=multiple imputation by chained equations, OR=odds ratio, PLCO=Prostate, Lung, Colon, Ovary Screening Trial, SD=standard deviation, TNFRSF11B=tumor necrosis factor receptor superfamily member 11B.

Enriched pathways were primarily related to immune and inflammatory signaling, including cytokine–cytokine receptor interactions, TNF and interleukin signaling, and immunoregulatory interactions between lymphoid and non-lymphoid cells. Additional enrichment was observed for cell adhesion molecule (CAM) and IgSF/CAM signaling, lysosomal and apoptotic processes, metabolic pathways, and hematopoietic cell lineage pathways. Together, these pathways indicate dysregulation of immune activation, cell–cell communication, and cellular turnover (**Figure 2**).

**Figure 2:**
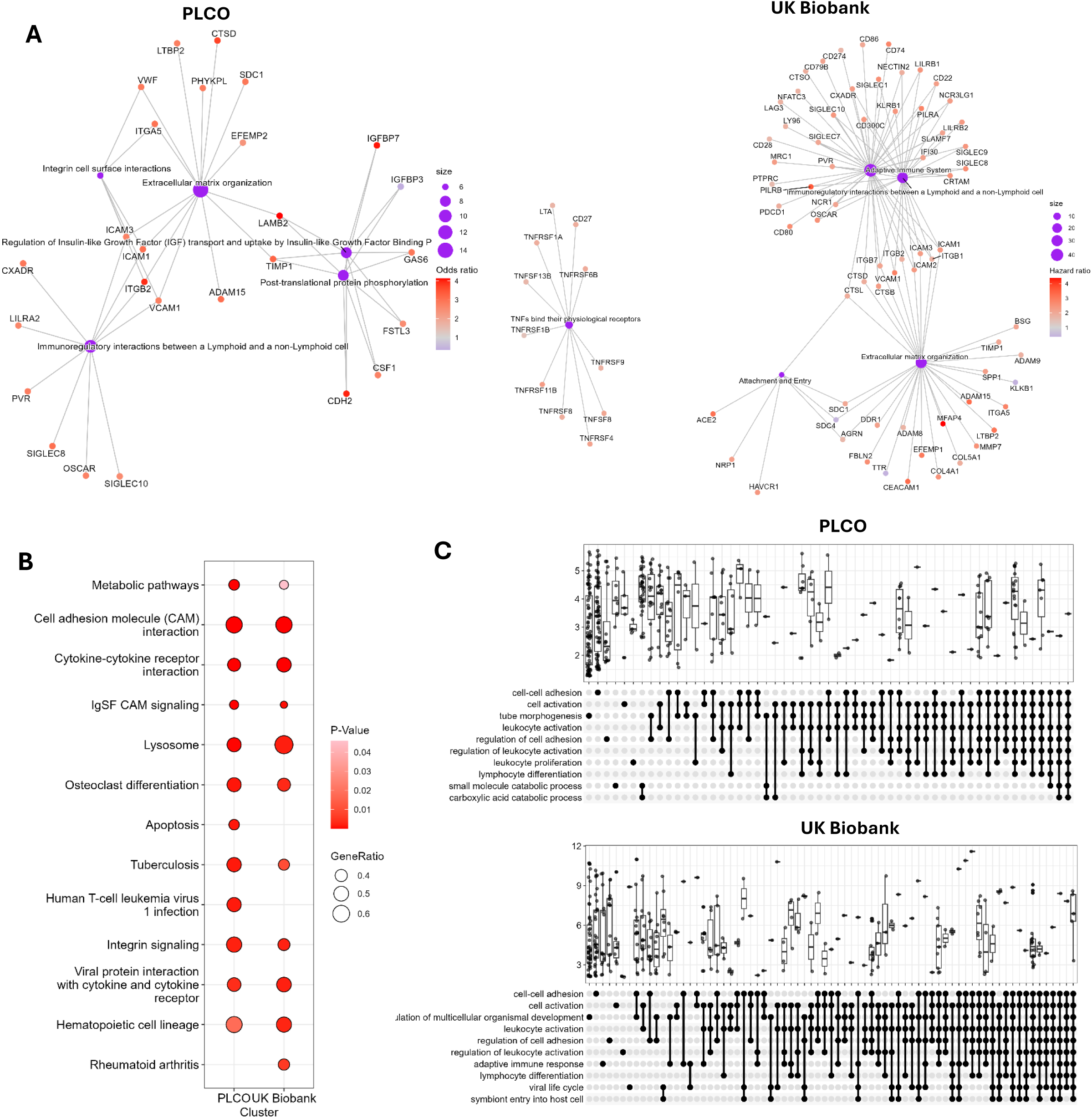
Pathway enrichment analyses of proteins associated with HCC risk. Over-representation analysis (panel A) was performed based on proteins significantly associated with HCC in comparison with the background set in each cohort. While gene set enrichment analysis (GSEA; panels B and D) used protein–HCC association rankings based on a composite score reflecting the magnitude, direction, and statistical significance of the log(RR). A. Reactome over-representation analysis of proteins significantly associated with HCC risk. The cnetplot shows the top enriched pathways and their connected proteins. Protein nodes are colored by effect size, and pathway nodes are shown in purple. B. Dot plot of KEGG GSEA results in PLCO and UK Biobank. Pathways are ordered by enrichment p-value in PLCO; dot size indicates the GeneRatio, and dot colour represents the enrichment p-value. D. UpSet plot of GO GSEA results. Each dot represents a leading-edge protein within an enriched GO term, and the y-axis displays the distribution of the ranking metric across pathways. Abbreviations: GSEA=gene set enrichment analyses, GO=Gene Ontology, HCC=hepatocellular carcinoma, KEGG= Kyoto Encyclopedia of Genes and Genomes, PLCO=Prostate, Lung, Colon, Ovary Screening Trial.

In sensitivity analyses excluding participants diagnosed ≤5 years of follow-up, 13 of the top 96 proteins (14%) available in both PLCO and UK Biobank had an attenuation in their log risk estimate greater than 5% compared with the primary analysis (**Supplementary Table 1** and **Supplementary Data**). We also compared associations of the 96 top proteins with associations reported in a published study from the Health Professionals Follow-up Study (HPFS) and Nurses’ Health Study (NHS) cohorts (SomaScan platform, 54 cases, 54 controls)^16^. Of the 96 top proteins, 37 were available for evaluation in this dataset, of which 18 demonstrated nominal (p < 0.05) and directionally consistent associations (**Supplementary Table 1**).

Overall, eleven proteins were Bonferroni significant in PLCO and UK Biobank, showed minimal temporal attenuation, and replicated in the HPFS/NHS cohort. The strongest associations were observed for complement component C7 (C7, OR per 1 SD = 3.61, 95% CI 2.17-6.02), scavenger receptor cysteine-rich type 1 protein M130 (CD163: 4.00, 2.29-6.98), and macrophage colony-stimulating factor 1 receptor (CSF1R: 3.07, 1.93-4.86) (**Supplementary Table 1**).

### 2.2. Multiprotein score to predict HCC occurrence

#### 2.2.1. Score development

In PLCO, the LASSO model identified eight proteins as collective predictors of HCC, listed in order of absolute effect size: adhesion G-protein coupled receptor G1 (ADGRG1), galectin-3-binding protein (LGALS3BP), thrombospondin-2 (THBS2), thymidine phosphorylase (TYMP), IGFBP3, ADAMTS-like protein 2 (ADAMTSL2), tetraspanin-8 (TSPAN8), and 4-hydroxyphenylpyruvate dioxygenase (HPD). Corresponding betas ranged between −0.06 to 0.32 (**Supplementary Table 2**). In 200 repeated analyses using random subsets of 50% of the data, ADGRG1 and LGALS3BP were selected in >94% of iterations (**Supplementary Figure 1**). In UK Biobank, all proteins were available in the protein risk score except HPD.

In both studies the score was approximately 4 SDs higher in future cases than controls (**Supplementary Figure 2**). Correlations between component proteins were moderate, with the highest correlation observed between THBS2 and ADAMTSL2 (r=0.71 in PLCO; r=0.53 in UK Biobank; **Supplementary Figure 3**).

The protein score was strongly associated with all established HCC risk factors in cancer-free controls, particularly HBV, HCV, alcoholic liver disease (ALD), MASLD, cirrhosis, diabetes, obesity, and age. For example, participants with HCV had a protein score 2.27 SDs (95% CI 1.92-2.61) higher than those without HCV (**Supplementary Figure 4**). Associations were broadly similar across the seven individual component proteins in UK Biobank (**Supplementary Table 3**).

#### 2.2.2. Risk prediction

In PLCO (training dataset), the score demonstrated high discriminatory power (AUC=0.90, 95% CI 0.86-0.94; sensitivity=0.77, and specificity=0.91), outperforming AFP (0.65, 0.58-0.72; sensitivity=0.62, specificity=0.64) (**Figure 3A**). Performance was similarly strong in the independent UK Biobank testing set (C-Index=0.92, 95% CI 0.84-0.97) compared with AFP (0.66, 0.55-0.76) (**Figure 3B**).

**Figure 3:**
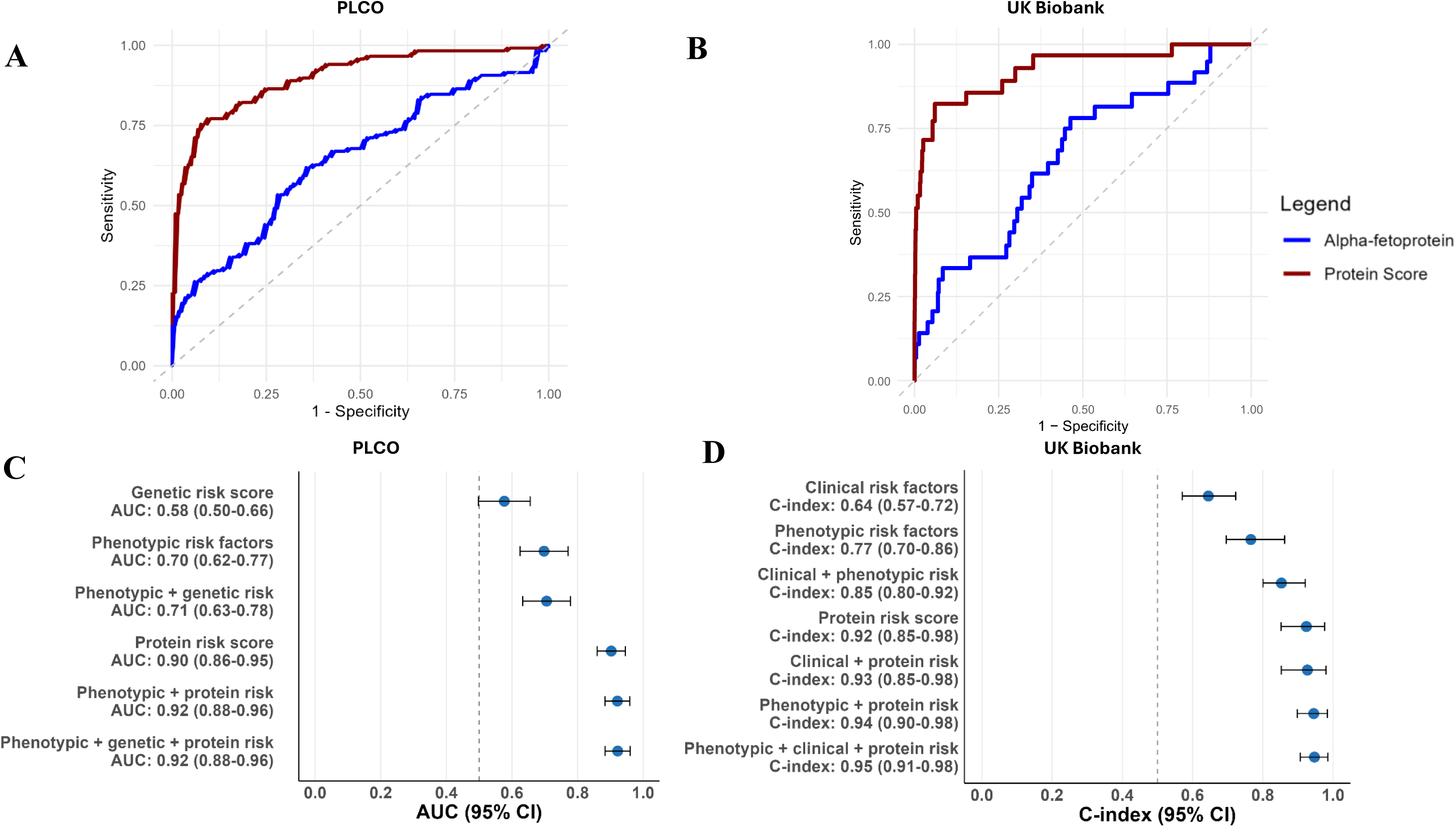
Multiprotein risk score for predicting HCC risk. **A:** Receiver operating characteristic curve for HCC in PLCO **B:** Receiver operating characteristic curve for HCC in UK Biobank at time 13.9 years (median total follow-up time) **C:** AUCs for HCC comparing multiprotein score, genetic risk score and phenotypic risk factors (age, sex, smoking, alcohol, BMI, and diabetes) performance in predicting HCC diagnosis. Data restricted to 96 cases and 105 controls with genetic and protein risk score data in PLCO **D:** C-indices for HCC comparing multiprotein score, clinical (cirrhosis, hepatitis B and hepatitis C prevalence at baseline) and phenotypic risk factors (age, sex, smoking, alcohol, BMI, and diabetes) performance in predicting HCC occurrence in UK Biobank Abbreviations: AUC=area under the curve, BMI=body mass index, CI=confidence interval, HCC=hepatocellular carcinoma, LASSO=least absolute shrinkage and selection operator, PLCO=Prostate, Lung, Colorectal and Ovarian Cancer Screening Trial

The protein risk score had higher discriminatory power than the genetic risk score (AUC=0.58, 0.50-0.66), phenotypic risk factors (AUC=0.70, 0.62-0.77; C-index=0.77, 0.70-0.86) and the presence of clinical risk factors (C-index=0.64, 0.57-0.72). Integrating genetic and established risk factors with the protein-based risk score marginally improved performance (PLCO AUC=0.92, 0.88-0.96; UK Biobank C-index: 0.95, 0.91-0.98) (**Figures 3C** and **3D**).

Predictive performance remained stable across follow-up periods examined in each study. In PLCO, the AUC was 0.90 (95% CI 0.84–0.95) for cases diagnosed within 10 years and 0.90 (0.84–0.96) for >10 years. In UK Biobank, the C-index was 0.91 (0.79–0.98) for ≤6.9 years of follow-up (median follow-up among cases) and 0.92 (0.84–0.99) for >6.9 years. The scores performed slightly worse in women, though the number of cases was small (PLCO: 22 cases, AUC=0.85, 0.73–0.97; UK Biobank: 8 cases, C-index=0.79, 0.57–0.96; **Table 2**).

**Table 2:** Associations of the multiprotein score for predicting hepatocellular carcinoma risk across subgroups.

| Subgroup | Stratum | PLCO |  | UK Biobank |  |
| --- | --- | --- | --- | --- | --- |
|  |  | Cases/<br>controls | AUC (95% CI) | N cases | C-index (95%<br>CI) |
| Time to diagnosis,<br>yrs | ≤Median* | 60 | 0.90 (0.84-0.95) | 15 | 0.91 (0.79-0.98) |
|  | >Median* | 58 | 0.90 (0.84-0.96) | 14 | 0.92 (0.84-0.99) |
| Death from liver<br>cancer | Yes | 80 | 0.89 (0.85-0.94) | 14 | 0.97 (0.92-0.99) |
| Grade | 1 | 13 | 0.94 (0.86-1.00) | - | - |
|  | 2 | 28 | 0.94 (0.87-1.00) | - | - |
|  | 3 | 13 | 0.77 (0.58-0.96) | - | - |
| Stage | Localized | 45 | 0.95 (0.90-1.00) | - | - |
|  | Regional | 20 | 0.89 (0.79-0.99) | - | - |
|  | Distant | 20 | 0.83 (0.70-0.96) | - | - |
| Sex | Men | 96/96 | 0.91 (0.87-0.95) | 21/19,518 | 0.97 (0.94-0.99) |
|  | Women | 22/22 | 0.85 (0.73-0.97) | 8/22,487 | 0.79 (0.57-0.95) |
| Age at blood<br>collection, yrs | ≤63.1 | 59/60 | 0.90 (0.84-0.96) | 16/30,006 | 0.91 (0.79-0.99) |
|  | >63.1 | 59/58 | 0.91 (0.85-0.96) | 13/11,999 | 0.94 (0.86-0.99) |
| Race | White | 95/95 | 0.90 (0.85-0.94) | 27/39,010 | 0.93 (0.85-0.98) |
|  | Non-white | 23/23 | 0.91 (0.82-1.00) | 2/2,790 | 0.82 (0.66-0.97) |
| BMI, kg/m<sup>2</sup> | <25 | 28/38 | 0.89 (0.79-0.98) | 8/13,623 | 0.96 (0.89-1.00) |
|  | 25-<30 | 50/58 | 0.89 (0.83-0.95) | 8/17,964 | 0.87 (0.65-1.00) |
|  | 30+ | 40/22 | 0.92 (0.85-0.98) | 12/10,209 | 0.93 (0.82-0.98) |
| Diabetes | No | 86/107 | 0.89 (0.84-0.94) | 23/39,505 | 0.91 (0.82-0.97) |
|  | Yes | 32/11 | 0.89 (0.78-1.00) | 6/2,315 | 0.96 (0.92-0.99) |

\*Median time to diagnosis in PLCO=10 years, UK Biobank=6.9 years
AUCs for hepatocellular carcinoma risk in PLCO were calculated from predicted probabilities generated by logistic regression models. For case-dependent characteristics controls were assigned the same value as their matched case. C-index for hepatocellular carcinoma risk in the UK Biobank was estimated using Cox models with time as the underlying time variable. CIs were derived using bootstrap with replacement (N iterations=1,000).
Abbreviations: AUC=area under the curve, BMI=body mass index, CI=confidence intervals, C-index=concordance index, PLCO=Prostate, Lung, Colorectal and Ovarian Cancer Screening Trial

ADGRG1 alone demonstrated high predictive performance (AUC=0.86, 0.81–0.91 and C-index=0.90, 0.81-0.96). Incrementally adding proteins to the score improved performance, but with minimal improvements observed beyond the top three proteins (**Supplementary Table 4**). The full score was notably associated with risk of developing liver conditions (ALD, MASLD, cirrhosis and fibrosis, viral hepatitis) after blood collection (e.g., HR of ALD per 1 SD=2.30, 95% CI 2.11-2.52) and all-cause mortality (1.24, 1.21-1.27), suggesting it may capture both subclinical and clinically evident liver dysfunction (**Supplementary Table 5**).

#### 2.2.3. Risk stratification in at-risk populations in UK Biobank

At-risk populations were defined as follows: (1) viral and non-viral liver conditions (current or past HBV/HCV infection or any liver condition, including cirrhosis, MASLD, and other liver disorders); (2) any non-viral liver condition; (3) cirrhosis and related fibrotic conditions (advanced liver disease); (4) non-viral liver conditions excluding advanced disease; and (5) viral liver condition (HBV/HCV). To maximize case numbers within at-risk categories, we additionally investigated associations with liver cancer. More information are provided in the **Supplementary Methods** and **Supplementary Table 6**.

Over a 7-year period, the annual risk of HCC was highest among participants with cirrhosis and related fibrotic conditions (1.0% per year), followed by HBV/HCV (0.4%) (**Supplementary Table 7**). In at-risk subsets, our protein risk score was strongly associated with HCC and liver cancer, demonstrating high discriminatory power, including in participants with cirrhosis (C-index for HCC=0.88, 95% CI 0.81-0.95) and HBV/HCV (0.93, 0.86-1.00), outperforming AFP (e.g., C-index for cirrhosis=0.67, 0.49-0.87; **Supplementary Table 8**).

When stratifying each at-risk population into lower- and higher-risk groups based on the protein score, the optimal threshold (using Youden’s Index) ranged from 2.1-3.5 SDs above the group-specific mean protein score (**Supplementary Table 9**). Using these cut-points, participants with cirrhosis and related conditions classified as higher-risk had an 8-year cumulative HCC risk of 17.6%, compared with 2.5% among those not classified as higher-risk, and corresponding liver cancer risks of 18.0% vs 3.3%. Similarly, among participants with HBV/HCV, 8-year cumulative HCC risk was 13.4% vs 1.0%, and liver cancer risk 15.2% vs 1.1% (**Figure 4**). The score also effectively stratified individuals with non-cirrhotic liver conditions (e.g., 8-year HCC risk 6.5% vs 0.44%) (**Supplementary Table 9**). Results were slightly stronger when analyses were restricted to individuals who were at-risk at baseline only, though case numbers were lower (e.g., cirrhosis 8-year HCC risk 25% vs 2.8%) (**Supplementary Table 10**).

**Figure 4:**
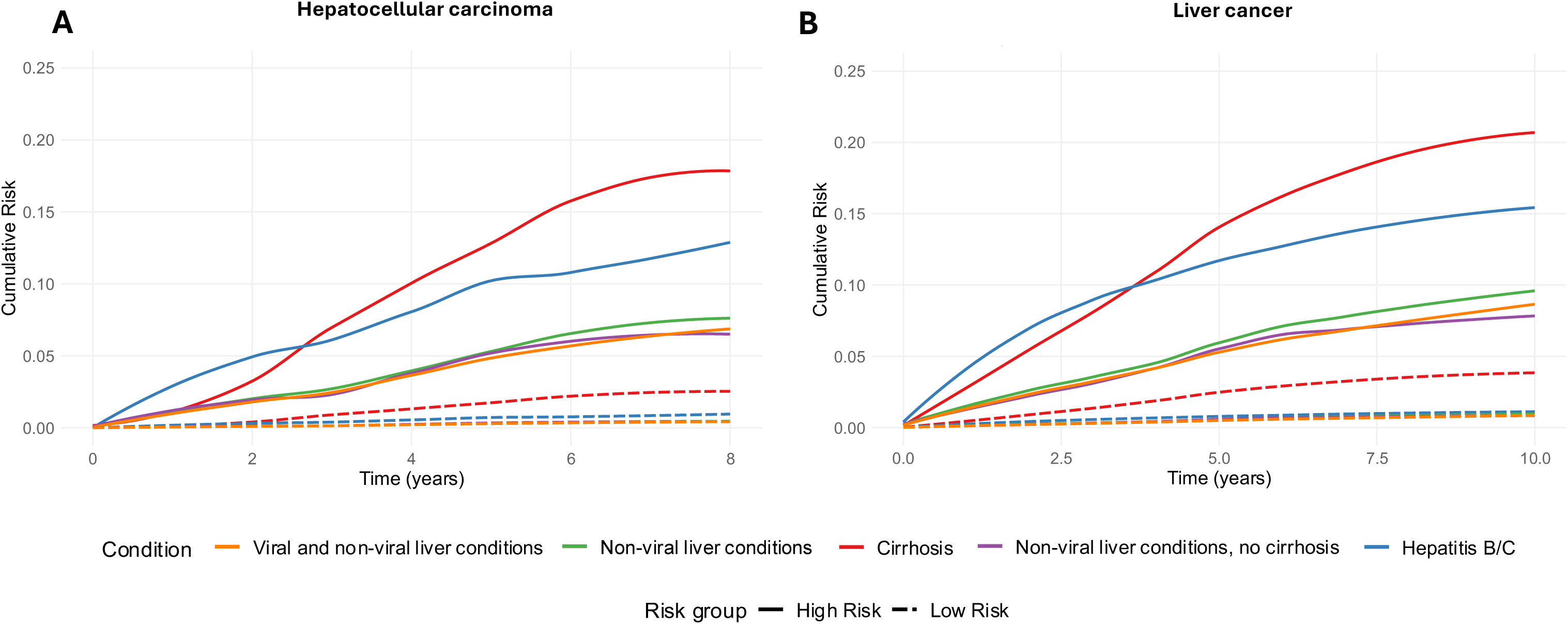
Multiprotein risk score for predicting risk in at-risk populations in the UK Biobank. **A:** Hepatocellular carcinoma **B:** Liver cancer Case numbers, C-index, and cut-points for each condition are available from **Supplementary Tables 8-9**. Associations based on Cox proportional hazards models with time on study as the underlying time variable. Follow-up time based on baseline blood collection for populations with prevalent disease (based on hospital records or responses to baseline interview) or first diagnosis with condition if occurred after study baseline (based on hospital admission records). Lines smoothed using locally estimating scatterplot smoothing curve. At-risk condition definitions available from **Supplementary Table 6**.

Score sensitivity ranged from 0.69-0.77 and specificity:0.74-0.87, outperforming AFP (sensitivity: 0.60-0.68; specificity:0.67-0.78) (**Supplementary Tables 9 and 11**). Sensitivity, specificity, and discriminatory performance were largely maintained over 8–10 years of follow-up (**Supplementary Table 12**). Although risk differences between score strata showed a modest attenuation after ∼6 years (**Figure 4**), the multiprotein score continued to outperform AFP across all follow-up time (**Supplementary Table 13**).

## 3. Discussion

In this analysis of over 4,000 circulating proteins, we identified 106 proteins significantly associated with HCC risk in PLCO, 97% of those available replicated in the UK Biobank. The associations of these individual proteins remained generally stable more than five years before diagnosis. Using a machine-learning approach, we developed a multiprotein risk score that demonstrated strong predictive performance across two independent cohorts and effectively stratified individuals with liver conditions into higher- and lower-risk groups up to eight years before HCC diagnosis.

The strong replication of protein associations across two independent cohorts was particularly notable. Additional evidence of replication from a published study in the HPFS and NHS cohorts further supports the validity and reproducibility of our findings. Where replication was not achieved, this may reflect differences in the proteomics platform and/or the limited sample size in the HPFS/NHS analytic set^16^. Many of the protein associations were also robust even for individuals diagnosed ≥5 years after baseline blood draw, indicating that many protein alterations likely preceded the onset of clinical disease. Among the proteins with the strongest evidence base were C7, CD163 and CSF1R, key markers of immune surveillance and inflammation. Previous clinical studies have reported these proteins as indicators of inflammatory burden, hepatic fibrosis, and progression toward HCC^22–27^. We also observed inverse associations for IGFBP-3, a major regulator of IGF-I signaling^28^. While inverse associations of IGFBP-3 have been documented across multiple cancers, including liver cancer, it remains unclear whether its effects on cancer are independent of IGF-I^29–32^. Further research is needed to clarify whether these proteins mediate associations of lifestyle and physiological factors (e.g., obesity) with HCC risk.

Current surveillance guidelines recommend bi-annual ultrasound (+/- AFP) for individuals with cirrhosis or chronic HBV^9,10^. However, the efficacy of this program is debated, given inconsistent evidence of reductions on cancer-related mortality, poor adherence, low sensitivity for early disease and among individuals with obesity^8,11,12,14,33–35^. Therefore, there is an urgent need for better tools to enhance risk prediction for early detection and risk stratification. Our protein risk score demonstrated strong predictive utility for HCC, with consistent performance years before diagnosis including among individuals with liver conditions.

ADGRG1 and LGALS3BP were the key contributors to our protein risk score. ADGRG1, a cell surface receptor, mediates cell adhesion and signaling, while LGALS3BP, a galectin-3–binding glycoprotein, regulates cell-cell interactions and immune responses. Both are frequently overexpressed across cancer types, including HCC^36–41^, and LGALS3BP has previously been proposed as a biomarker for HCC detection and prognosis^42–44^. Their oncogenic roles likely involve altered cell migration, angiogenesis, immune evasion, and apoptosis^45–49^, potentially through interaction with the transforming growth factor-β pathway^41,48,50^, which drives fibrosis, cirrhosis, and HCC development^50–53^. Although both proteins were linked to incident liver conditions, the protein score further stratified individuals by HCC risk, suggesting it reflects processes beyond liver damage and captures early neoplastic transformation.

If validated in an independent clinical population, a multi-protein risk score could surpass AFP as a biomarker or serve as a complementary tool within existing surveillance protocols^54^. For instance, it could help identify individuals at the highest risk individuals who might benefit from more sensitive imaging, such as magnetic resonance imaging, which has higher early-stage sensitivity (∼84%) but remains too resource-intensive for routine use^11,35,55^. For lower-risk individuals, longer surveillance intervals could be explored to optimize resource use and support compliance. For individuals with non-cirrhotic liver conditions, such as the 30-40% of MALFD-related HCC cases that occur without cirrhosis^56,57^, current guidelines do not recommend surveillance as HCC risk is lower (0.1-1.3 cases per 1,000 patient-years)^56^. A simplified and affordable protein panel might provide a minimally invasive approach to identify those higher-risk individuals who could benefit from targeted screening. Overall, by supporting earlier detection and individualized risk stratification, a proteomics-based approach could substantially lessen healthcare burden and contribute to reductions in HCC mortality. Clinical validation in at-risk populations remains critical to advance these findings toward practice.

Strengths of our investigation include the long follow-up period (up to 21 years), replication of findings in studies nested within two independent prospective cohorts, and evidence of consistency based on a third study using an aptamer-based platform^16^. Another key strength is the breadth of proteins, encompassing low-abundance and secretome proteins, many of which have not been previously investigated. The Olink antibody-based platforms are highly specific, enabling precise protein identification^58,59^. The routine linkage to hospitalization and national health registries in the UK Biobank facilitates identification and long-term follow-up of individuals with liver conditions, minimizing attrition and misclassification biases^60^.

This study has several limitations. Proteins were measured using different platforms—Explore 3072 in UK Biobank and HT platform in PLCO, therefore ∼25% of proteins in PLCO were unavailable for replication. Additionally, UK Biobank used EDTA-plasma samples, while PLCO used serum, though protein levels are likely highly correlated between these sample types and associations were highly consistent^61^. The small number of HCC cases limited the power to examine associations stratified by follow-up time, and by factors such as sex and race. Participants were predominantly non-Hispanic White and healthier than the general population, which may reduce generalizability of risk estimates and highlights the need for additional data in women and non-White populations. The prevalence of cirrhosis and HBV was also low in both cohorts. Validation of risk stratification in at-risk populations (e.g., clinicaltrials.gov NCT00001971, NCT00006164) using absolute protein concentrations is an important next stage^62,63^. Our protein score was compared to AFP levels as measured using the Olink platform, whereas clinical settings typically use ELISA with absolute cut-points (e.g., 20 ng/mL)^35^, and it is unknown whether at-risk participants engaged in HCC surveillance. To address smaller sample sizes in at-risk subsets, participants could enter the at-risk group after study baseline, which reduced the average follow-up duration.

In conclusion, we identified 93 proteins consistently associated with HCC across two independent cohorts, many remaining stable at least five years before diagnosis, implicating inflammation and growth-factor signaling in disease etiology. Our multiprotein score showed strong discriminatory power and effectively stratified individuals with cirrhosis or HBV/HCV into higher- and lower-risk groups up to eight years of follow-up. To translate these findings into practice, future studies in at-risk populations should focus on developing cost-effective assays and evaluating whether proteomic risk stratification can improve early detection and reduce liver cancer mortality.

## 4. Methods

This study followed the Strengthening the Reporting of Observational Studies in Epidemiology (STROBE) reporting guideline for cohort studies^64^.

### 4.1. PLCO

#### 4.1.1. Study design

The PLCO study is a multicenter randomized cancer screening trial comprising ∼155,000 participants (ages 55-74) with no prior prostate, lung, colorectal, or ovarian cancer^65,66^. Our analysis was nested within the 77,000 screening-arm participants who provided non-fasting blood samples. All participants provided written informed consent and the study was approved by the Institutional Review Boards at the National Cancer Institute and the 10 screening centers.

We included 119 HCC incident cases (International Classification of Diseases, 10^th^ Edition [ICD-10]: C22, ICD-O-3: 8170–8175), diagnosed through 2017 and 119 matched cancer-free controls (**Supplementary Figure 5**). Cases and controls were individually matched by age at blood draw, sex, race/ethnicity, study year of blood draw and year of randomization. Median time from blood collection to HCC diagnosis was 9.7 years (maximum=21.4). One sample was later excluded due to quality control criteria related to protein measurements (*Section 4.1.2*.). Further study design details are provided in the **Supplementary Methods**.

#### 4.1.2. Protein assessment

Proteins in serum samples were measured using the Olink Explore HT platform, which utilizes proximity extension assays, where antibody pairs conjugated to complementary oligonucleotides hybridize upon target binding, enabling relative quantification. The platform quantified 5,419 assays, representing 5,415 distinct proteins with a few proteins measured by multiple assays. Normalized Protein eXpression (NPX) values were derived by normalizing raw assay counts to the Extension Control for each sample and block (defined by Olink based on protein abundance and sample dilution), log_2_ transformed, and intensity normalized.

We excluded one sample because 81% of the assays did not pass QC, and a further 1,055 assays flagged by Olink as “failed” or “warning”. For proteins with values below the limit of detection (LOD), provided by Olink, we imputed to half of the LOD^67^.

To evaluate platform performance across the 5,419 assays we included sample replicates, freeze-thawed samples and one-year temporal repeats (see **Supplementary Methods**). Median intra- and inter-plate intraclass correlations (ICCs) were 0.91 (IQR 0.71-0.98) and 0.88 (0.66-0.95), with CVs of 6.1% (0-10.6%) and 11.7% (8.4-16.7%), respectively. The median 1-year temporal ICC was 0.72 (0.51-0.87), and the mean absolute percentage difference due to freeze-thaw was 14.4% (9.91–20.0%), slightly above the 10.2% (5.8-16.5%) for technical repeats (**Supplementary Figure 6**).

We excluded 1,416 proteins with >95% of values below the LOD, leaving a final analytic set of 4,003 proteins. There was no strong evidence of measurement outliers based on principle component analyses or quality control metrics (ICCs and CVs); therefore, no additional proteins were excluded.

### 4.2. UK Biobank

#### 4.2.1. Study design

The UK Biobank is a prospective cohort of 502,150 individuals ages 37-73 years, recruited through the UK National Health Service from 2006-2010. Of 9.2 million invited, 5.5% attended baseline assessments at 22 centers across England, Wales, and Scotland^68^. The UK Biobank study was approved by the North West Multi-Centre Research Ethics Committee (Reference Number 21/NW/0157). The proteomics subset comprised randomly selected 46,595 individuals, 6,376 selected by the UK Biobank Pharma Proteomics Project (UKB-PPP), and 1,268 from a COVID-19 imaging study^69^.

Cancer and death data were obtained through record linkage with national registries. Follow-up was defined as the end of follow-up (May 31, 2023 in England; December 31, 2016 in Wales; and September 30, 2023 in Scotland), first malignant cancer diagnosis, death, or loss to follow-up, whichever occurred first. HCC diagnoses (ICD-10: C22, ICD-O-3:8170-8175) were identified in 36 participants after a median 13.9 years of follow-up.

#### 4.2.2. Protein assessment

Proteins were measured using the Olink Explore 1536 and Explore Expansion platforms in ∼54,000 participants at study baseline. EDTA-plasma samples were analyzed at the Olink Analyses Service, Uppsala, Sweden using the Olink Explore platforms, which captured 2,923 distinct proteins targeted by 2,941 assays. Proteins were log_2_ transformed, intensity normalized and harmonized using bridging samples^69^.

The UKB-PPP QC process included internal controls, outlier detection via principal component analysis and NPX distribution checks. Samples with QC warnings, assay failures, or suspected swaps were removed, leaving 52,995 participants with valid protein measurement (median CV 6.7%)^69^.

We excluded 2,812 participants with prevalent cancer based on cancer registries and one participant with no follow-up time (**Supplementary Figure 7**). We also excluded two proteins which were available in less than 10 cases (GLIPR1 and NPM1). In total, our analytic population included 50,182 participants and 2,921 proteins.

#### 4.2.3. At-risk populations

Participants at-risk for developing HCC were identified using baseline interview and hospital admission record. We categorized individuals into groups, capturing a broad range of severity, HCC risk, and etiology: (1) viral and non-viral liver conditions (current or past HBV/HCV diagnosis [ICD-10: B16, B17.0-B17.1, B18.0-B18.2, B18.8-B18.9, B19.1-B19.2] or any liver condition [K70-K77]); (2) any non-viral liver condition (K70-K77 only); (3) non-viral liver conditions excluding advanced disease (K70.0-K70.2, K70.4-K70.9, K71-K73.9, K74.7-K77); (4) cirrhosis, hepatic fibrosis and sclerosis (K70.3, K74.0-K74.6), representing more severe liver pathology; and (5) viral liver conditions (current or past HBV/HCV infection). Full definitions, including ICD-9 and self-report codes, and censoring dates are provided in the **Supplementary Methods** and **Supplementary Table 6**.

Both prevalent conditions at baseline and incident diagnoses during follow-up were included, provided they occurred before a cancer diagnosis. To maximize statistical power in these smaller “at-risk” groups, we also investigated associations with liver cancer (C22). Follow-up began at baseline for individuals with prevalent liver conditions and at the date of first diagnosis for those who developed a liver condition after baseline, and continued until cancer diagnosis, death, loss to follow-up, or censoring date (**Supplementary Methods**).

### 4.3. Statistical analysis

#### 4.3.1. Protein imputation and standardization

In PLCO, imputation was unnecessary due to minimal missing protein data (0.05%). In UK Biobank, proteins were imputed using random forests (500 trees, max iterations=7) for participants missing <20% of proteins and proteins missing <20% of participants (N participants=41,839, N proteins=2,919), with age and sex as covariates^70,71^. Standardization in PLCO was based on control distribution (mean=0, SD=1), while in UK Biobank, it was based on the whole sample.

#### 4.3.2. Protein associations with HCC

Protein–HCC associations were estimated using conditional logistic regression conditioned on matching factors in PLCO, and Cox regression in UK Biobank with follow-up time as the underlying time scale. Models were adjusted for age, smoking, alcohol consumption, education, BMI, and diabetes status in both cohorts, with additional adjustments for sex and race in UK Biobank (matching factors in PLCO). Missing covariates were imputed using multiple imputation by chained equations^72^. Associations were Bonferroni-corrected for multiple testing (PLCO:4,003 tests; UK Biobank: 2,921). Associations were meta-analyzed using a fixed-effects model.

To identify biological pathways associated with circulating proteins and HCC risk, enrichment analyses was performed using the Reactome, Kyoto Encyclopedia of Genes and Genomes (KEGG), and Gene Ontology (GO) databases, which offer complementary perspectives on biological processes and signaling pathways to capture a more comprehensive view of underlying pathways^73^. Reactome pathway enrichment analysis was conducted on proteins significantly associated with HCC risk within each cohort compared to all the proteins measured in each cohort. These results were visualized using network plots, illustrating the connections between proteins and enriched pathways. For KEGG and GO pathway analysis, proteins were ranked based on the direction and magnitude of the log risk estimates and p-values with HCC risk, and enrichment was quantified using Fast Gene Set Enrichment Analysis^74^ using the clusterProfiler R package^75^.

In sensitivity analyses we excluded participants diagnosed within the first 5 years of follow-up and compared risk estimates to those from the primary analysis. We also compared our reported associations with the largest previously published study of HCC in the HPFS and NHS cohorts, using the SomaScan platform (N proteins = 1,305, N cases = 54 and N controls = 54)^16^.

#### 4.3.3. Protein risk score

##### 4.3.3.1. Score development

We integrated multiple protein biomarkers into an optimized risk score in PLCO using least absolute shrinkage and selection operator (LASSO) regression^76^. Following a previously described approach^18^, models were trained using 3-fold cross-validation repeated 5 times, and performance was assessed using Cohen’s Kappa, with final models selected based on the optimal lambda values. To evaluate consistency of protein selection, we generated 200 random subsamples of the PLCO dataset, each containing 50% of the data. The score was then computed in UK Biobank using the beta coefficients derived from PLCO.

Recognizing that elevated scores may reflect broader liver disease burden and overall health risk beyond HCC, we investigated associations with incident liver conditions and all-cause mortality using Cox models. Associations with sociodemographic, lifestyle and clinical risk factors were examined using linear regression models. These analyses were not conducted in PLCO due to the limited number of controls and absence of liver disease among them.

##### 4.3.3.2. Score performance

In PLCO, score performance was evaluated using AUC, and the C-index (AUC for time to event data) in UK Biobank, with 95% CIs estimated via bootstrapping (1,000 iterations). Protein risk score was compared to AFP, established lifestyle/health risk factors (age, smoking, alcohol, BMI, and diabetes), clinical risk factors (HBV, HCV, and cirrhosis) and genetic risk^77^. We also examined score performance by time to diagnosis, liver cancer death, stage and grade (PLCO only), sex, age, race, BMI, and diabetes.

##### 4.3.3.3. At-risk populations

In UK Biobank, we evaluated the score’s performance in at-risk populations and its ability to stratify participants by HCC risk. Optimal cutpoints for each at-risk population was determined by dividing the score into 100 percentiles and iteratively fitting Cox models to maximize Youden’s Index at 3 years (median time-to-diagnosis in at-risk groups). Using these cut-points, we stratified participants into higher- and lower-risk groups and calculated the mean predicted cumulative risk for each (see **Supplementary Methods**). Additionally, we estimated the annual risk of developing HCC in each risk group by dividing the mean predicted cumulative risk over the study duration by the number of years in the study. In sensitivity analyses, we also restricted at-risk individuals to those who were prevalent at baseline only.

## Data sharing statement

We do not have permission to share the data directly. However, all bona fide researchers can apply to use the PLCO (https://cdas.cancer.gov/plco/) and UK Biobank (https://www.ukbiobank.ac.uk/register-apply/) resources for health-related research that is in the public interest.

## Code availability

Code used for the analyses is available at: https://github.com/elliewatts90/proteins_hepatocellular_carcinoma.

## Disclaimer

The contributions of the NIH authors are considered Works of the United States Government. The findings and conclusions presented in this paper are those of the author(s) and do not necessarily reflect the views of the NIH or the U.S. Department of Health and Human Services.

## Supporting information

Government coversheet

## Acknowledgements

We thank all of the participants, researchers, and support staff who made the study possible. This work utilized the computational resources of the NIH HPC Biowulf cluster (http://hpc.nih.gov). Cancer incidence data for PLCO have been provided by the Alabama Statewide Cancer Registry, Arizona Cancer Registry, Colorado Central Cancer Registry, District of Columbia Cancer Registry, Georgia Cancer Registry, Hawaii Cancer Registry, Cancer Data Registry of Idaho, Maryland Cancer Registry, Michigan Cancer Surveillance Program, Minnesota Cancer Surveillance System, Missouri Cancer Registry, Nevada Central Cancer Registry, Ohio Cancer Incidence Surveillance System, Pennsylvania Cancer Registry, Texas Cancer Registry, Utah Cancer Registry, Virginia Cancer Registry, and Wisconsin Cancer Reporting System. All are supported in part by funds from the Center for Disease Control and Prevention, National Program for Central Registries, local states or by the National Cancer Institute, Surveillance, Epidemiology, and End Results program. The results reported here and the conclusions derived are the sole responsibility of the authors.

## Additional Information

This research was conducted using PLCO (Project ID: 2023-0061) and the UK Biobank Resource under application number 92005.

