## Supplementary Methods, Figures, and Tables for "Proteomic analysis of hepatocellular carcinoma etiology and risk stratification in two prospective studies"

#### Supplementary Figures

### Supplementary Methods

#### I) Prostate, Lung, Colorectal, and Ovarian Cancer Screening Trial

##### Design and sample selection

The PLCO study is a population-based multicenter randomized screening trial of people aged 55-74 years at baseline with no history of prostate, lung, colorectal, or ovarian cancer[1,2]. This study was approved by institutional review boards at the US National Cancer Institute and the 10 study centers and all participants signed informed consent.

The eligible population for our study comprised 77,443 participants of the screening arm who completed baseline questionnaires on smoking, diet, and other health-related characteristics[3]. All variables collected at study baseline were self-reported. Diet was assessed using a dietary history questionnaire, where participants reported how often, on average, they consumed food in the last year.

Serum samples were collected at baseline in the screening arm and annually for five years, except year 3. For all hepatocellular carcinoma (HCC) cases and controls, we prioritized baseline serum samples that had not undergone prior freeze-thaw cycles if available, if not, we selected samples from the next available time-point without freeze-thaw. Cases and controls were individually matched by age at blood draw, sex, race/ethnicity, study year of blood draw and year of randomization. In total, 119 cases and 119 controls selected: 85 cases from baseline, 8 from year 1, 22 from year 2, 2 from year 4, and 2 from year 5. Controls were selected from the same study year as their matched case.

##### Genetic risk score

Genome-wide genotyping was performed using OncoArray, Omni Express, Omni2.5 M, and the Illumina Global Screening Array across different individuals. Genotype imputation was conducted using the TOPMed reference panel via the Michigan Imputation Server[3]. The genetic risk score for HCC was derived from independent single nucleotide polymorphisms (SNP) significantly associated with HCC in the largest genome-wide association study[4], with each allele weighted by its beta coefficient. One SNP (rs58489806) was unavailable in PLCO. The genetic risk score was standardized (mean=0, standard deviation [SD]=1) based on the distribution among the controls.

##### Olink Explore HT platform evaluation

For quality assessment, we included six sets of four replicate samples, 12 freeze-thawed replicates, and 12 one-year temporal repeats, all randomly selected from study controls. To improve sample diversity beyond the HCC cases and controls, which were predominantly White men, we additionally included blood samples from 8 cancer-free White women, 28 Black women, and 28 Black men. Although these samples were not analyzed in our nested case-control study, they were included in our platform assessment to estimate between-subject variance.

Measurements below the limit of detection (LOD) were imputed to half the LOD. Intraclass correlation coefficients (ICCs) were estimated using linear mixed-effects models, which partitioned the variance into between-participant and within-participant components. The ICC represents the proportion of total variance attributable to between-participant variability, rather than measurement error. Due to the high proportion of imputed protein data (many values were below the limit of detection), between-subject variance was estimated using the entire study population rather than being restricted to quality control replicates. Intra-plate ICCs were estimated by calculating the mean ICC for each plate and protein, then calculating the median across all proteins (median ICC=0.91, IQR=0.71-0.98). Inter-plate ICCs were derived by calculating the mean replicate protein sample within each plate, followed by calculating the median ICC across all proteins (median inter-plate ICC=0.88, IQR=0.66-0.95).

Coefficients of variation (CVs) were calculated based on replicate samples only, with measurement values squared to reverse the log2 transformation. The median intra-plate CV was 6.1% (IQR=0.0–10.6%), while the median inter-plate CV was 11.7% (IQR=8.4–16.7%).

ICCs for temporal and freeze-thaw samples were calculated using similar methods as described above, with between-subject variance estimated from the whole study population. Temporal analyses excluded freeze-thaw samples, and freeze-thaw analyses excluded temporal samples. The median 1-year temporal ICC was 0.72 and 0.80 for freeze-thaw samples. The mean absolute percentage difference was 14.4% (IQR=9.91–20.0%) for freeze-thawed samples and 10.2% (IQR=5.8–16.5%) for technical repeats (**Supplementary Figure 6**).

#### Variable coding

Associations between each protein and HCC risk were estimated using conditional logistic regression models conditioned on the matching factors. Models were further adjusted for age (continuous), smoking status (never, former, current), alcohol consumption (none, >0–≤1, >1–≤2, >2 drinks/day), coffee intake (continuous), educational attainment (up to high school, some post-secondary training, college graduate or higher), body mass index (<25, 25–<30, ≥30 kg/m²), and diabetes (no, yes). Missing covariate data were imputed using multiple imputation by chained equations (MICE).

### II) UK Biobank

#### Design and sample selection

The UK Biobank is a prospective cohort of 502,150 individuals ages (37-73) recruited through the UK National Health Service from 2006-2010. Of 9.2 million invited, 5.5% attended baseline assessments at 22 centers across England, Wales, and Scotland[5]. The UK Biobank study was approved by the North West Multi-Centre Research Ethics Committee (Reference Number 21/NW/0157). The proteomics subset comprised randomly selected 46,595 individuals, 6,376 selected by the UK Biobank Pharma Proteomics Project (UKB-PPP), and 1,268 from a COVID-19 imaging study[6].

At the assessment center, trained staff measured height and weight, and a nurse-led interview was used to record non-cancer comorbidities. Lifestyle, sociodemographic, and selected health factors (e.g., diabetes) were collected using a touchscreen questionnaire, and dietary intake was assessed from self-reported average consumption.

#### Variable coding

We used Cox proportional hazards regression models, with follow-up time as the underlying time scale. Models were adjusted for age (continuous), sex (men, women), race (White, all other racial groups), alcohol consumption (<1, ≥1 drinks/day), coffee intake (continuous, cups/day), smoking status (never, former, current), educational attainment (no college or equivalent professional qualification, college graduate or higher), BMI (<25, 25–<30, ≥30 kg/m²), and diabetes (self-report: no, yes). Coffee consumption was truncated at the 90th percentile, and a standard drink was defined as containing 14 g of alcohol for comparability with U.S. population measures. Missing covariates were imputed using MICE.

#### Identification of at-risk populations

We identified individuals at-risk of developing liver cancer using responses from the baseline interview and hospital admissions data (codes available from **Supplementary Table 6**). The definition of at-risk groups was intentionally broad, encompassing individuals with any diagnosis of liver disease or viral hepatitis (B and C). This approach ensured sufficient numbers for stable estimates, particularly given the smaller sample sizes in at-risk categories. More specific definitions, such as cirrhosis or viral hepatitis B, were also examined to provide refined analyses in groups typically monitored for HCC[7,8]. At-risk groups included conditions either prevalent at study baseline or diagnosed during follow-up, providing there was no prior recorded cancer diagnosis (excluding non-melanoma skin cancer [C44]).

Hospital admission data includes any patient who is admitted to the hospital and occupies a bed (both for emergency and planned admissions) but does not include outpatients or accident and emergency (unless patient is subsequently admitted)[9]. Diagnoses of liver conditions were obtained through record linkage (England: Hospital Episode Statistics for England, censoring date 31st March 2023, Scotland: Scottish Morbidity Record, 31st August 2022, and Wales: Patient Episode Database for Wales, 31st May 2022). More information for these data is available from: https://biobank.ndph.ox.ac.uk/showcase/showcase/docs/HospitalEpisodeStatistics.pdf and published elsewhere[9]. Death records were obtained from NHS England (England and Wales) and NHS Central Register, National Records of Scotland.

Cancer diagnoses were identified using cancer registry data up to the registry-specific censoring dates (May 31, 2023 in England; December 31, 2016 in Wales; and September 30, 2023 in Scotland). For HCC, maximal censoring dates were based on country-specific cancer registry dates, as diagnosis requires histological confirmation. To maximize statistical power for analyses within at-risk groups, liver cancer cases were additionally included as an outcome, drawing on both cancer registry and death records. In Wales, cancer registry follow-up is shorter than hospital admission follow-up; therefore, to extend follow-up time, we incorporated liver cancer cases identified from hospital records when they occurred after the cancer registry censoring date.

For individuals with prevalent liver conditions (identified through self-report or hospital admission records prior to baseline), follow-up began at the date of their baseline assessment and continued until cancer diagnosis, death, loss to follow-up, or the cancer registry censoring date, whichever occurred first. For individuals who developed a liver condition after baseline, follow-up began on the date the condition was first diagnosed.

Analyses were restricted to liver-related conditions with at least five HCC cases.

#### Score performance in at-risk populations

Cox proportional hazards models were used to assess protein score performance in at-risk populations. For participants with a prevalent liver condition at baseline, the baseline date was used as the entry time in the model. For those who developed a liver condition during follow-up without a prior cancer diagnosis, the entry time was set to the date of the first recorded occurrence of the condition. These models were used to estimate cumulative baseline hazard values, similar to methods described previously[10].

In brief, the cumulative baseline hazard function ($H_0$ at time $t$) is given as:

$$H_0(t) = \int_0^t \lambda_0(u) du$$

where $\lambda_0(t)$ is the baseline hazard function at time t. The baseline survival probability (which represents the probability of remaining free from liver cancer) at time t is given as:

$$S_0(t) = \exp(-H_0(t)).$$

Individual predicted risks were then derived using the baseline survival probability and the linear predictor $\eta$ from the Cox model:

$$\eta = XB,$$

where $X$ are the covariates and $B$ are the predicted cumulative risk of HCC at time t:

$$R(t) = 1 - S_0(\mathrm{t})^{\exp(\eta)}.$$

Participants were stratified into high- and lower-risk groups based on an optimal risk score cutpoint and mean cumulative risks in the high risk (positive predictive value, PPV), low risk (complement of the negative predictive value, cNPV), and overall risk groups were calculated.

Sensitivity at time *t*, measures the proportion of true positives detected among cases and is calculated as:

$$Sensitivity(t) = \frac{P(M+, D+)}{P(D+)},$$

where P(M+, D+) is the fraction of the population in the high-risk group multiplied by the PPV and P(D+) is the average population-level risk after 8-years for HCC and 10-years for liver cancer. As some individuals in the UK Biobank are right-censored before the specified time point, the average population-level risks are estimated using Cox models.

Specificity at time *t*, measures the proportion of true negatives detected among non-cases and is calculated as:

$$Specificity(t) = \frac{P(M-, D-)}{P(D-)},$$

where P(M-, D-) is the fraction of the population in the low-risk group multiplied by 1-cNPV, and P(D-) is 1-P(D+).

Condition-specific optimal cutpoints were determined by dividing the protein risk score into 100 percentiles and identifying the value that maximized Youden's Index (sensitivity + specificity - 1) at 3 years of follow-up, which corresponds to the approximate median time to diagnosis in the high-risk population.

#### Software and packages

Analyses were conducted in R version 4.3.1, using R packages: mice (3.18.0)[11], survival (3.8.3)[12], caret (7.0.1)[13], missRanger (4.4.3)[14], glmnet (4.1.10)[15], and clusterProfiler (4.14.6)[16].

### Supplementary Figures

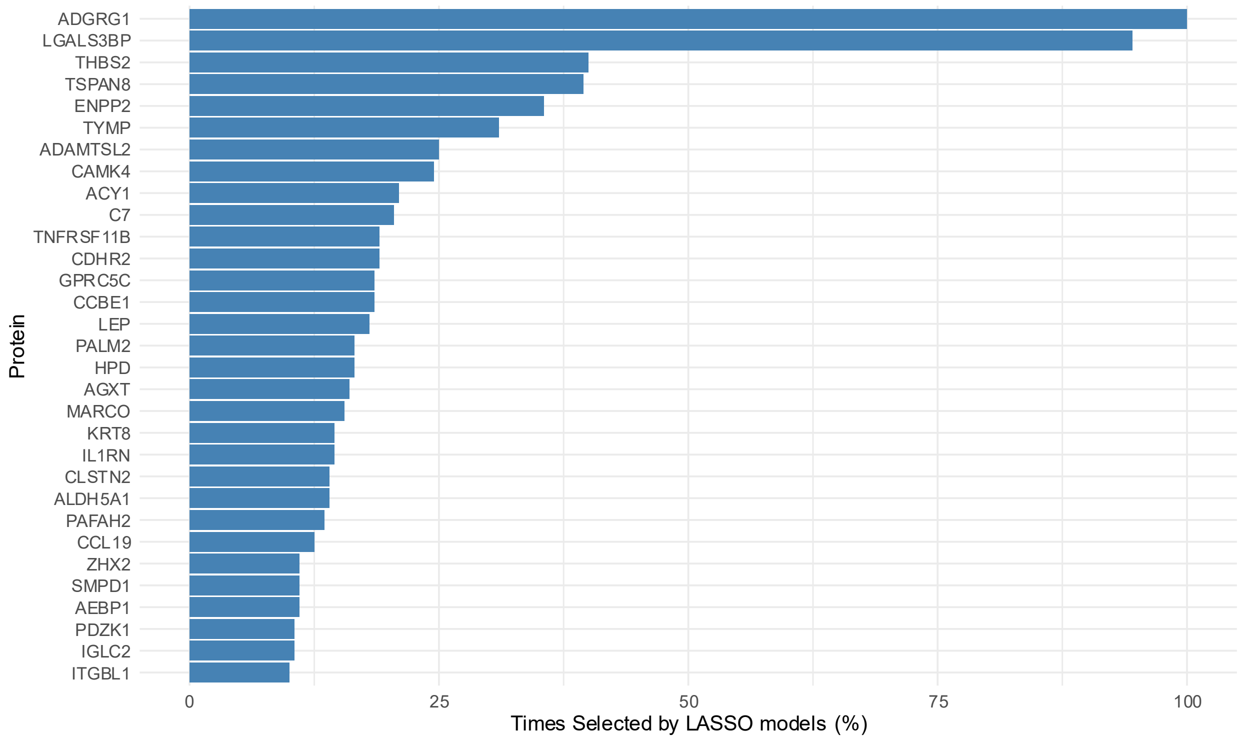

**Supplementary Figure 1: Frequency of protein selection using LASSO Models in PLCO**

The frequency of protein selection was assessed using LASSO regression with 3-fold cross-validation repeated 5 times, applied to 200 random subsets of the data, each comprising 50% of the total dataset.

Abbreviations: LASSO=least absolute shrinkage and selection operator, PLCO=Prostate, Lung, Colorectal and Ovarian Cancer Screening Trial

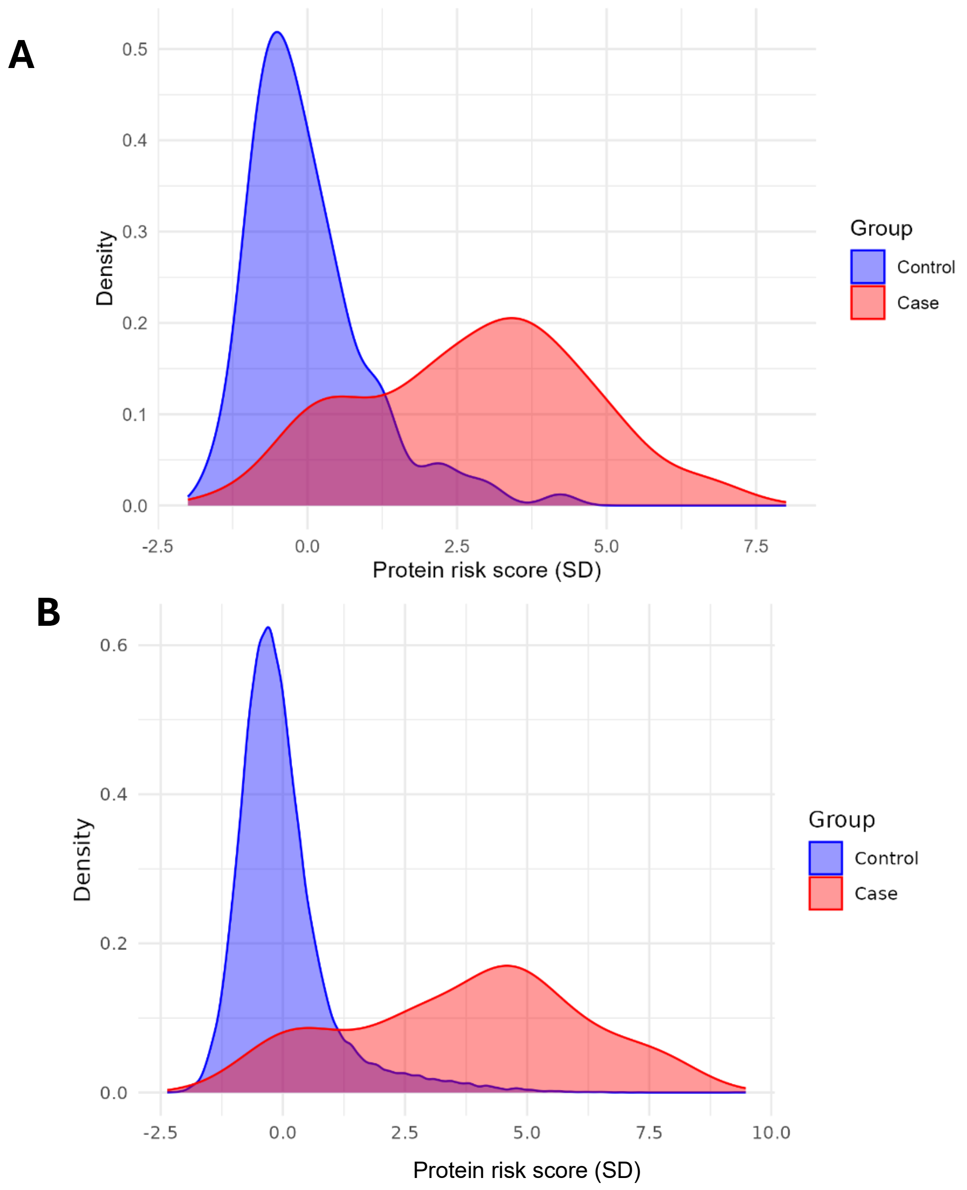

**Supplementary Figure 2: Distribution of the protein risk score in future HCC cases and controls**

**A: PLCO**
**B: UK Biobank**
Abbreviations: HCC=hepatocellular carcinoma, PLCO=Prostate, Lung, Colorectal and Ovarian Cancer Screening Trial, SD=standard deviation.

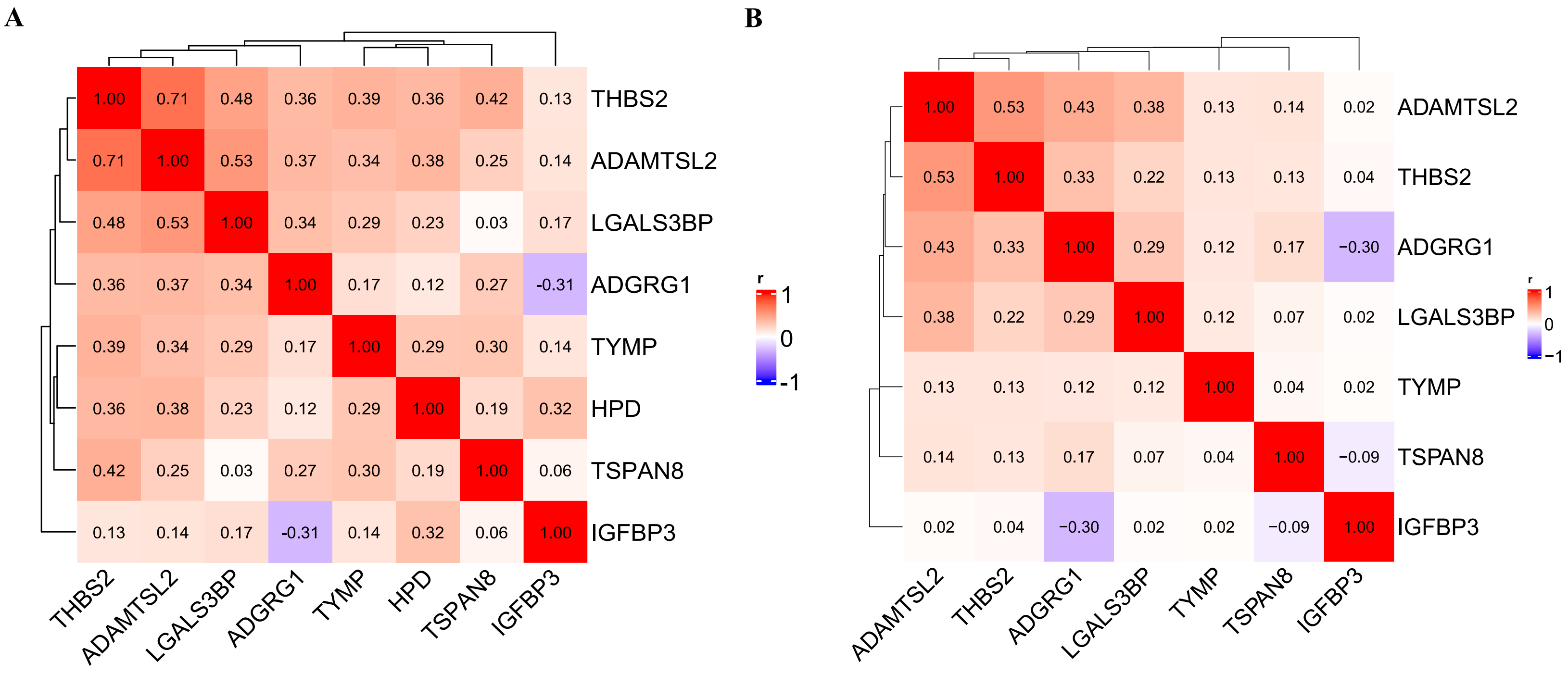

**Supplementary Figure 3: Pearson correlations between proteins retained in LASSO model in controls**

**A) PLCO**
**B) UK Biobank**

Abbreviations: HCC=hepatocellular carcinoma, LASSO=least absolute shrinkage and selection operator, PLCO=Prostate, Lung, Colorectal and Ovarian Cancer Screening Trial

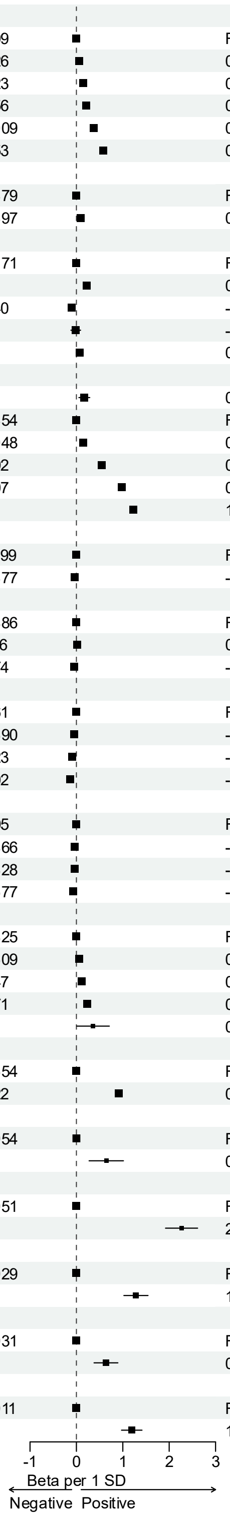

| Variable | N | Beta per 1 SD (95% CI) | P-value |
|---|---|---|---|
| Age | | | |
| <45 | 4,409 | Ref | |
| 45-49 | 5,426 | 0.06 (0.02, 0.10) | $7.0\times10^{-4}$ |
| 50-54 | 6,223 | 0.14 (0.11, 0.18) | $2.2\times10^{-16}$ |
| 55-59 | 7,456 | 0.21 (0.18, 0.25) | $4.8\times10^{-37}$ |
| 60-64 | 10,009 | 0.37 (0.34, 0.40) | $3.6\times10^{-116}$ |
| 65+ | 8,453 | 0.58 (0.55, 0.61) | $1.4\times10^{-264}$ |
| Sex | | | |
| Women | 22,479 | Ref | |
| Men | 19,497 | 0.09 (0.07, 0.11) | $5.1\times10^{-21}$ |
| Race | | | |
| White | 39,171 | Ref | |
| Asian | 965 | 0.22 (0.16, 0.27) | $1.4\times10^{-13}$ |
| Black | 1,040 | -0.11 (-0.16, -0.05) | $1.8\times10^{-4}$ |
| Mixed race | 276 | -0.01 (-0.12, 0.09) | 0.789 |
| Other | 524 | 0.07 (-0.01, 0.15) | 0.071 |
| BMI | | | |
| <18.5 | 219 | 0.17 (0.05, 0.29) | 0.004 |
| 18.5-24.9 | 13,454 | Ref | |
| 25-29.9 | 18,048 | 0.14 (0.12, 0.16) | $3.3\times10^{-46}$ |
| 30-34.9 | 7,292 | 0.54 (0.52, 0.57) | $<2\times10^{-308}$ |
| 35-39.9 | 2,107 | 0.97 (0.93, 1.01) | $<2\times10^{-308}$ |
| 40+ | 856 | 1.22 (1.16, 1.28) | $1.1\times10^{-308}$ |
| Education | | | |
| <College | 11,499 | Ref | |
| College+ | 30,477 | -0.04 (-0.06, -0.01) | 0.008 |
| Alcohol (drinks/d) | | | |
| < 1 | 25,886 | Ref | |
| >2 | 7,116 | 0.02 (-0.01, 0.04) | 0.149 |
| 1-2 | 8,974 | -0.05 (-0.07, -0.03) | $4.2\times10^{-6}$ |
| Coffee (cups/d) | | | |
| 0 | 9,461 | Ref | |
| 1-2 | 19,390 | -0.05 (-0.07, -0.03) | $1.2\times10^{-5}$ |
| 3-4 | 8,523 | -0.09 (-0.12, -0.07) | $9.9\times10^{-12}$ |
| 5+ | 4,602 | -0.13 (-0.17, -0.10) | $1.7\times10^{-15}$ |
| Tea (cups/d) | | | |
| 0 | 6,105 | Ref | |
| 1-2 | 10,866 | -0.04 (-0.06, -0.01) | 0.011 |
| 3-4 | 12,328 | -0.04 (-0.07, -0.01) | 0.003 |
| 5+ | 12,677 | -0.07 (-0.10, -0.04) | $1.3\times10^{-6}$ |
| Smoking status | | | |
| Never | 22,825 | Ref | |
| Previous | 14,609 | 0.06 (0.04, 0.08) | $2.2\times10^{-9}$ |
| Current, <15 cig/d | 2,747 | 0.11 (0.07, 0.14) | $1.1\times10^{-9}$ |
| Current, 15+ cig/d | 1,771 | 0.23 (0.18, 0.27) | $1.5\times10^{-24}$ |
| Current unknown | 24 | 0.36 (0.01, 0.71) | 0.045 |
| Diabetes | | | |
| No | 39,654 | Ref | |
| Yes | 2,322 | 0.91 (0.88, 0.95) | $<2\times10^{-308}$ |
| Hepatitis B | | | |
| No | 41,954 | Ref | |
| Yes | 22 | 0.64 (0.27, 1.01) | $6.7\times10^{-4}$ |
| Hepatitis C | | | |
| No | 41,951 | Ref | |
| Yes | 25 | 2.27 (1.92, 2.61) | $1.2\times10^{-37}$ |
| ALD | | | |
| No | 41,929 | Ref | |
| Yes | 47 | 1.28 (1.02, 1.54) | $5.0\times10^{-22}$ |
| MASLD | | | |
| No | 41,931 | Ref | |
| Yes | 45 | 0.64 (0.38, 0.89) | $1.2\times10^{-6}$ |
| Cirrhosis | | | |
| No | 41,911 | Ref | |
| Yes | 65 | 1.19 (0.97, 1.41) | $3.7\times10^{-26}$ |

**Supplementary Figure 4: Associations of the multiprotein score with HCC risk factors in participants did not develop HCC during the study period in UK Biobank**

Associations estimated using linear models based on variables at study baseline. All associations are mutually adjusted for the other variables. Associations for each individual protein are available from **Supplementary Table 3**.

Abbreviations: ALD=alcoholic liver disease, BMI=body mass index, CI=confidence interval, HCC=hepatocellular carcinoma, MASLD=metabolic dysfunction-associated steatotic liver disease, SD=standard deviation.

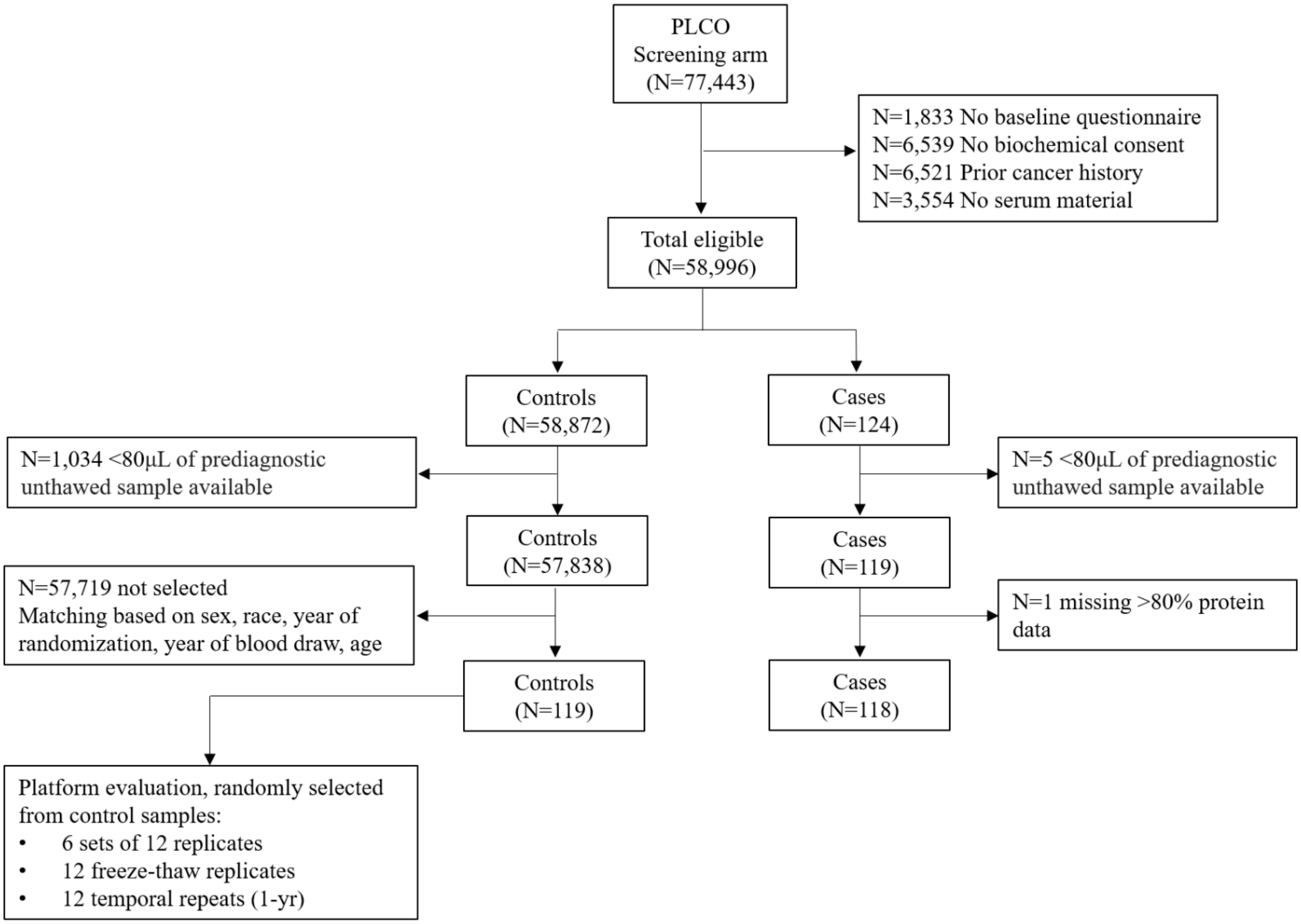

**Supplementary Figure 5: Participant selection criteria in PLCO**

For analyses conditioned on matching variables (conditional logistic regression models), only complete matched case–control sets were included (118 cases matched to 118 controls). For analyses not conditioned on matching factors, complete sets were not required; thus, 118 cases and 119 controls were included.

Abbreviations: PLCO=Prostate, Lung, Colorectal and Ovarian Cancer Screening Trial

**A**

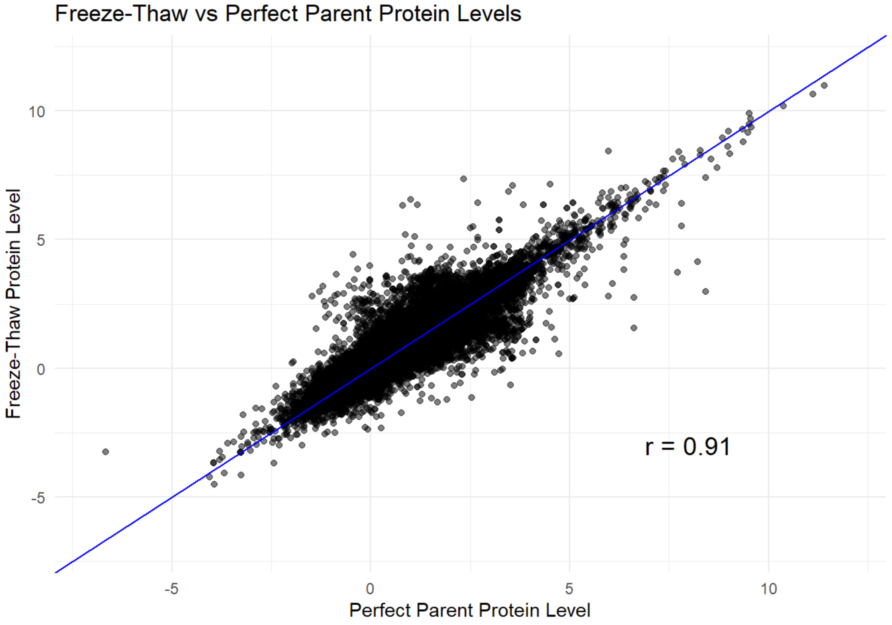

**B**

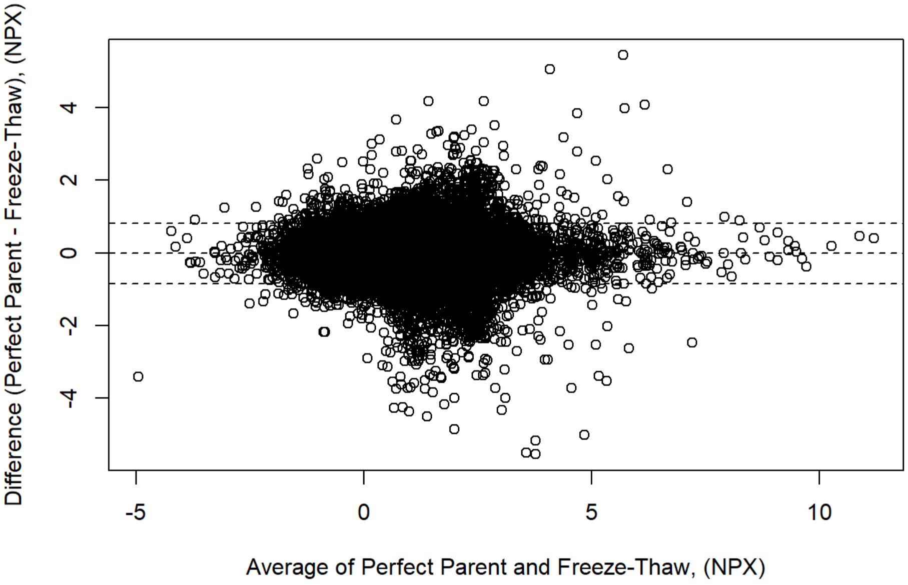

**Supplementary Figure 6: Comparison of samples with one prior freeze-thaw with replicate samples with no prior freeze-thaw**

**A**: Correlation between measurements
**B**: Bland-Altman plot showing the difference in protein measurements between perfect parent samples and freeze-thawed samples as a function of their average. The dashed lines represent the mean difference (center line) and ±1.96 standard deviations (outer lines), illustrating the limits of agreement.
The analysis was based on 12 perfect parent serum samples with no prior freeze-thaw cycles and their corresponding 12 replicate samples that had undergone one freeze-thaw cycle. These samples were randomly selected from the control group.

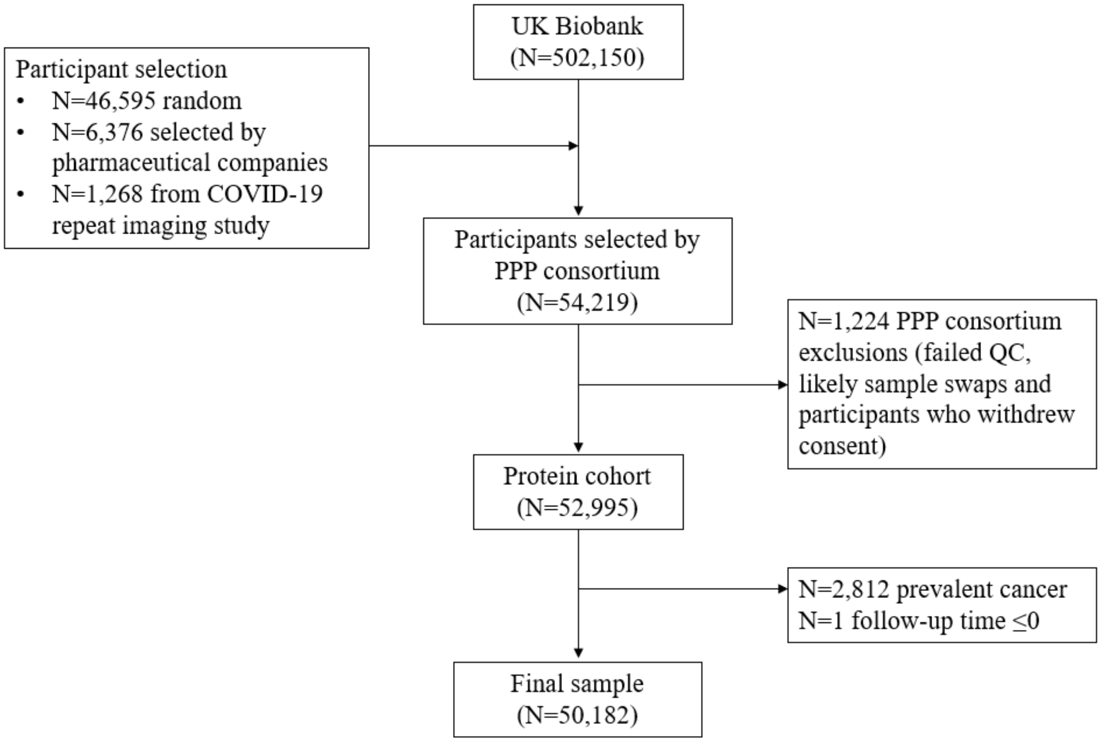

**Supplementary Figure 7: Participant selection criteria in UK Biobank**

### Supplementary Tables

**Supplementary Table 1: Robust protein associations with hepatocellular carcinoma in PLCO and UK Biobank**

| | PLCO | | | UK Biobank | | | | | |
|---|---|---|---|---|---|---|---|---|---|
| Protein | Case/ Control | OR per 1 SD (95% CI) | P-value | Cases/ Total | HR per 1 SD (95% CI) | P-value | Bonferroni significant in both cohorts | No large temporal attenuation * | Consistent HPFS/ NHS† |
| C7 | 118/118 | 3.61 (2.17, 6.02) | 2E-06 | 31/42,845 | 3.06 (2.58, 3.64) | 1E-10 | 1 | 1 | 1 |
| CD163 | 118/118 | 4.00 (2.29, 6.98) | 3E-06 | 35/49,701 | 3.15 (2.52, 3.95) | 6E-10 | 1 | 1 | 1 |
| CSF1R | 118/118 | 3.07 (1.93, 4.86) | 5E-06 | 31/42,865 | 1.91 (1.66, 2.20) | 2E-08 | 1 | 1 | 1 |
| IL18BP | 118/118 | 2.58 (1.75, 3.81) | 5E-06 | 35/49,701 | 2.58 (2.18, 3.06) | 1E-10 | 1 | 1 | 1 |
| VCAM1 | 118/118 | 2.75 (1.81, 4.16) | 5E-06 | 35/49,701 | 2.87 (2.46, 3.35) | 3E-12 | 1 | 1 | 1 |
| IGFBP7 | 118/118 | 4.06 (2.27, 7.27) | 6E-06 | 35/49,746 | 1.77 (1.61, 1.94) | 3E-10 | 1 | 1 | 1 |
| CSF1 | 118/118 | 2.70 (1.78, 4.10) | 7E-06 | 35/49,595 | 2.43 (1.90, 3.09) | 2E-07 | 1 | 1 | 1 |
| KYNU | 118/118 | 3.08 (1.91, 4.95) | 9E-06 | 34/49,591 | 2.25 (1.68, 3.02) | 1E-05 | 1 | 1 | 1 |
| PIGR | 118/118 | 4.38 (2.34, 8.20) | 9E-06 | 35/49,446 | 2.63 (2.17, 3.20) | 1E-09 | 1 | 1 | 1 |
| CTSD | 118/118 | 3.68 (2.12, 6.40) | 9E-06 | 35/49,411 | 1.92 (1.69, 2.18) | 6E-10 | 1 | 1 | 1 |
| TNFRSF1B | 118/118 | 2.75 (1.78, 4.24) | 1E-05 | 35/49,453 | 1.43 (1.29, 1.59) | 6E-07 | 1 | 1 | 1 |
| TNFRSF11B | 118/118 | 2.86 (1.91, 4.30) | 1E-06 | 35/49,593 | 2.32 (1.95, 2.76) | 6E-09 | 1 | 1 | 0 |
| KRT18 | 118/118 | 2.83 (1.87, 4.30) | 3E-06 | 36/49,335 | 2.58 (2.05, 3.23) | 1E-08 | 1 | 1 | 0 |
| ICAM1 | 118/118 | 3.04 (1.93, 4.78) | 4E-06 | 35/49,697 | 2.38 (2.05, 2.76) | 5E-11 | 1 | 1 | 0 |
| PLAU | 118/118 | 3.48 (2.09, 5.79) | 5E-06 | 35/49,453 | 1.40 (1.30, 1.51) | 9E-09 | 1 | 1 | 0 |
| ICAM3 | 118/118 | 2.69 (1.79, 4.05) | 5E-06 | 35/49,746 | 2.30 (1.85, 2.85) | 7E-08 | 1 | 1 | 0 |
| ENG | 118/118 | 3.62 (2.13, 6.16) | 5E-06 | 35/49,746 | 2.06 (1.78, 2.38) | 3E-09 | 1 | 1 | 0 |
| ERBB2 | 118/118 | 4.27 (2.34, 7.78) | 6E-06 | 36/49,471 | 2.67 (2.14, 3.34) | 6E-09 | 1 | 1 | 0 |
| ADGRE2 | 118/118 | 2.57 (1.73, 3.80) | 6E-06 | 35/49,595 | 2.61 (2.20, 3.11) | 2E-10 | 1 | 1 | 0 |
| CD80 | 118/118 | 4.90 (2.52, 9.50) | 7E-06 | 31/42,773 | 2.60 (2.18, 3.10) | 3E-09 | 1 | 1 | 0 |
| MSR1 | 118/118 | 4.10 (2.27, 7.39) | 7E-06 | 35/49,548 | 3.70 (2.89, 4.76) | 4E-10 | 1 | 1 | 0 |
| FSTL3 | 118/118 | 2.51 (1.70, 3.69) | 8E-06 | 35/49,593 | 2.56 (2.08, 3.16) | 6E-09 | 1 | 1 | 0 |
| FAP | 118/118 | 2.89 (1.84, 4.54) | 1E-05 | 35/49,746 | 2.51 (1.94, 3.23) | 2E-07 | 1 | 1 | 0 |
| PDGFRA | 118/118 | 2.29 (1.61, 3.27) | 1E-05 | 35/49,618 | 2.35 (2.01, 2.75) | 2E-10 | 1 | 1 | 0 |
| CDH6 | 118/118 | 3.00 (1.88, 4.80) | 1E-05 | 35/49,618 | 2.33 (2.02, 2.68) | 7E-11 | 1 | 1 | 0 |
| CDH2 | 118/118 | 3.98 (2.22, 7.17) | 1E-05 | 35/49,618 | 2.76 (2.07, 3.68) | 4E-07 | 1 | 1 | 0 |
| IL1RL1 | 118/118 | 2.86 (1.82, 4.49) | 1E-05 | 35/49,743 | 2.63 (1.94, 3.57) | 2E-06 | 1 | 1 | 0 |
| IL4R | 118/118 | 2.30 (1.61, 3.29) | 1E-05 | 34/49,461 | 2.12 (1.84, 2.45) | 8E-10 | 1 | 1 | 0 |
| CDCP1 | 118/118 | 3.76 (2.25, 6.28) | 1E-06 | 34/49,335 | 2.66 (2.09, 3.37) | 4E-08 | 1 | 1 | N/A |
| CLSTN2 | 118/118 | 2.44 (1.72, 3.46) | 2E-06 | 34/49,591 | 2.28 (1.95, 2.68) | 8E-10 | 1 | 1 | N/A |
| SLAMF8 | 118/118 | 3.75 (2.22, 6.34) | 2E-06 | 34/49,123 | 2.66 (2.23, 3.16) | 2E-10 | 1 | 1 | N/A |
| ITGB2 | 118/118 | 3.93 (2.27, 6.78) | 3E-06 | 35/49,734 | 1.83 (1.61, 2.09) | 3E-09 | 1 | 1 | N/A |
| SPON2 | 118/118 | 3.23 (2.02, 5.16) | 3E-06 | 35/49,618 | 3.38 (2.69, 4.24) | 3E-10 | 1 | 1 | N/A |
| CEACAM1 | 118/118 | 4.72 (2.53, 8.81) | 3E-06 | 36/49,395 | 3.29 (2.71, 4.00) | 1E-11 | 1 | 1 | N/A |
| NFASC | 118/118 | 3.84 (2.24, 6.59) | 3E-06 | 34/49,591 | 4.49 (3.45, 5.86) | 2E-10 | 1 | 1 | N/A |
| PALM2 | 118/118 | 2.97 (1.92, 4.61) | 3E-06 | 30/42,663 | 1.88 (1.57, 2.25) | 2E-06 | 1 | 1 | N/A |
| MME | 118/118 | 3.61 (2.15, 6.05) | 3E-06 | 36/49,335 | 3.82 (2.74, 5.31) | 2E-08 | 1 | 1 | N/A |
| SIGLEC10 | 118/118 | 2.53 (1.74, 3.69) | 4E-06 | 34/49,461 | 2.41 (2.03, 2.86) | 8E-10 | 1 | 1 | N/A |
| SCLY | 118/118 | 2.64 (1.78, 3.92) | 4E-06 | 35/49,472 | 2.30 (1.73, 3.06) | 5E-06 | 1 | 1 | N/A |
| ITGA5 | 118/118 | 2.98 (1.92, 4.64) | 4E-06 | 35/49,453 | 2.82 (2.31, 3.44) | 4E-10 | 1 | 1 | N/A |
| ADGRE1 | 118/118 | 3.07 (1.95, 4.84) | 4E-06 | 30/42,666 | 2.51 (2.01, 3.14) | 2E-07 | 1 | 1 | N/A |
| SULT2A1 | 118/118 | 2.53 (1.73, 3.70) | 5E-06 | 34/49,461 | 1.82 (1.55, 2.13) | 2E-07 | 1 | 1 | N/A |
| PLA2G15 | 118/118 | 3.77 (2.19, 6.49) | 5E-06 | 36/49,471 | 2.50 (2.05, 3.05) | 2E-09 | 1 | 1 | N/A |
| IFI30 | 118/118 | 3.74 (2.17, 6.43) | 5E-06 | 30/42,649 | 2.19 (1.77, 2.72) | 8E-07 | 1 | 1 | N/A |
| ADGRE5 | 118/118 | 3.34 (2.03, 5.50) | 5E-06 | 35/49,743 | 1.81 (1.63, 2.02) | 1E-10 | 1 | 1 | N/A |
| PRCP | 118/118 | 3.17 (1.97, 5.09) | 6E-06 | 35/49,595 | 1.81 (1.56, 2.11) | 7E-08 | 1 | 1 | N/A |
| PTPRH | 118/118 | 3.65 (2.14, 6.23) | 6E-06 | 30/42,666 | 2.15 (1.80, 2.57) | 1E-07 | 1 | 1 | N/A |
| ADAM15 | 118/118 | 3.06 (1.92, 4.86) | 6E-06 | 34/49,370 | 3.25 (2.44, 4.31) | 3E-08 | 1 | 1 | N/A |
| GRPEL1 | 118/118 | 3.55 (2.09, 6.01) | 6E-06 | 36/49,335 | 2.21 (1.85, 2.63) | 3E-09 | 1 | 1 | N/A |
| CREG1 | 118/118 | 3.11 (1.94, 5.00) | 7E-06 | 36/49,393 | 1.88 (1.52, 2.33) | 3E-06 | 1 | 1 | N/A |
| TSPAN8 | 118/118 | 5.48 (2.71, 11.10) | 7E-06 | 30/42,666 | 3.47 (2.53, 4.75) | 3E-07 | 1 | 1 | N/A |
| ANGPTL2 | 118/118 | 3.19 (1.97, 5.18) | 7E-06 | 35/49,592 | 2.43 (1.83, 3.24) | 2E-06 | 1 | 1 | N/A |
| GAS6 | 118/118 | 2.90 (1.86, 4.52) | 7E-06 | 35/49,741 | 3.32 (2.58, 4.27) | 2E-09 | 1 | 1 | N/A |
| CD74 | 118/118 | 2.59 (1.74, 3.86) | 7E-06 | 35/49,548 | 2.77 (2.21, 3.46) | 5E-09 | 1 | 1 | N/A |
| CD28 | 118/118 | 2.99 (1.89, 4.74) | 7E-06 | 34/49,123 | 1.67 (1.49, 1.89) | 2E-08 | 1 | 1 | N/A |
| IL32 | 118/118 | 2.99 (1.89, 4.73) | 8E-06 | 34/49,461 | 1.96 (1.72, 2.23) | 1E-09 | 1 | 1 | N/A |
| ADA2 | 118/118 | 3.07 (1.92, 4.91) | 8E-06 | 35/49,696 | 2.78 (2.36, 3.28) | 1E-11 | 1 | 1 | N/A |

| | | | | | | | | | |
|---|---|---|---|---|---|---|---|---|---|
| CXADR | 118/118 | 2.90 (1.85, 4.54) | 8E-06 | 34/49,461 | 2.28 (1.91, 2.72) | 5E-09 | 1 | 1 | N/A |
| AGXT | 118/118 | 3.78 (2.15, 6.64) | 9E-06 | 35/49,618 | 3.05 (2.31, 4.04) | 5E-08 | 1 | 1 | N/A |
| MILR1 | 118/118 | 2.30 (1.62, 3.28) | 9E-06 | 34/49,255 | 2.30 (1.86, 2.85) | 8E-08 | 1 | 1 | N/A |
| ADGRG1 | 118/118 | 3.98 (2.22, 7.16) | 9E-06 | 36/49,335 | 2.94 (2.38, 3.63) | 4E-10 | 1 | 1 | N/A |
| C1QTNF1 | 118/118 | 4.08 (2.25, 7.42) | 9E-06 | 35/49,701 | 1.86 (1.57, 2.21) | 2E-07 | 1 | 1 | N/A |
| SMPD1 | 118/118 | 2.54 (1.71, 3.78) | 9E-06 | 35/49,465 | 2.10 (1.65, 2.67) | 3E-06 | 1 | 1 | N/A |
| LTBP2 | 118/118 | 2.76 (1.79, 4.24) | 1E-05 | 35/49,746 | 3.11 (2.55, 3.79) | 7E-11 | 1 | 1 | N/A |
| PTS | 118/118 | 2.38 (1.64, 3.45) | 1E-05 | 34/49,114 | 2.06 (1.70, 2.49) | 2E-07 | 1 | 1 | N/A |
| C19ORF12 | 118/118 | 2.71 (1.77, 4.15) | 1E-05 | 34/49,118 | 1.64 (1.41, 1.91) | 1E-06 | 1 | 1 | N/A |
| SIGLEC8 | 118/118 | 3.05 (1.89, 4.92) | 1E-05 | 31/42,695 | 2.58 (1.95, 3.41) | 2E-06 | 1 | 1 | N/A |
| PVR | 118/118 | 2.80 (1.80, 4.37) | 1E-05 | 35/49,548 | 2.13 (1.72, 2.63) | 3E-07 | 1 | 1 | N/A |
| SRPX | 118/118 | 2.73 (1.77, 4.22) | 1E-05 | 31/42,773 | 2.53 (2.11, 3.02) | 3E-09 | 1 | 1 | N/A |
| PTPRK | 118/118 | 2.50 (1.68, 3.70) | 1E-05 | 30/42,666 | 1.45 (1.29, 1.62) | 3E-06 | 1 | 1 | N/A |
| HTRA2 | 118/118 | 3.62 (2.11, 6.19) | 7E-06 | 36/49,471 | 1.43 (1.03, 2.00) | 4E-02 | 0 | 1 | 1 |
| ECHS1 | 118/118 | 2.42 (1.70, 3.46) | 3E-06 | 31/42,695 | 1.47 (1.03, 2.10) | 3E-02 | 0 | 1 | N/A |
| IGSF9 | 118/118 | 2.72 (1.80, 4.13) | 6E-06 | 31/42,773 | 2.52 (1.79, 3.56) | 3E-05 | 0 | 1 | N/A |
| GSTA1 | 118/118 | 2.60 (1.73, 3.92) | 1E-05 | 35/49,544 | 2.10 (1.54, 2.85) | 6E-05 | 0 | 1 | N/A |
| PCDHB15 | 118/118 | 2.24 (1.59, 3.15) | 1E-05 | 30/42,668 | 1.64 (1.37, 1.98) | 3E-05 | 0 | 1 | N/A |
| PCBD1 | 118/118 | 2.82 (1.81, 4.38) | 1E-05 | 31/42,809 | 1.82 (1.41, 2.36) | 1E-04 | 0 | 1 | N/A |
| LILRA2 | 118/118 | 2.63 (1.74, 3.97) | 1E-05 | 35/49,453 | 2.46 (1.68, 3.59) | 7E-05 | 0 | 1 | N/A |
| KHK | 118/118 | 2.13 (1.54, 2.94) | 1E-05 | 31/42,695 | 1.78 (1.20, 2.64) | 7E-03 | 0 | 1 | N/A |
| PHYKPL | 118/118 | 2.83 (1.81, 4.41) | 1E-05 | 31/42,809 | 1.80 (1.31, 2.48) | 1E-03 | 0 | 1 | N/A |
| KCTD5 | 118/118 | 3.22 (1.95, 5.31) | 1E-05 | 30/42,657 | 1.50 (1.28, 1.76) | 7E-05 | 0 | 1 | N/A |
| GBP1 | 118/118 | 2.65 (1.74, 4.03) | 1E-05 | 30/42,663 | 1.80 (1.39, 2.32) | 2E-04 | 0 | 1 | N/A |
| CHI3L1 | 118/118 | 3.82 (2.25, 6.49) | 2E-06 | 35/49,593 | 2.59 (2.06, 3.26) | 2E-08 | 1 | 0 | 1 |
| IGFBP3 | 118/118 | 0.35 (0.23, 0.54) | 3E-06 | 35/49,701 | 0.43 (0.36, 0.51) | 3E-09 | 1 | 0 | 1 |
| ACY1 | 118/118 | 3.91 (2.24, 6.83) | 4E-06 | 35/49,618 | 2.71 (2.01, 3.65) | 7E-07 | 1 | 0 | 1 |
| GDF15 | 118/118 | 3.25 (1.99, 5.32) | 7E-06 | 35/49,746 | 2.06 (1.76, 2.41) | 4E-09 | 1 | 0 | 1 |
| TIMP1 | 118/118 | 3.22 (1.95, 5.34) | 1E-05 | 35/49,685 | 2.05 (1.81, 2.32) | 8E-11 | 1 | 0 | 1 |
| NRCAM | 118/118 | 2.37 (1.64, 3.43) | 1E-05 | 35/49,618 | 1.95 (1.64, 2.33) | 9E-08 | 1 | 0 | 0 |
| NRP1 | 118/118 | 3.35 (1.99, 5.64) | 1E-05 | 35/49,704 | 2.22 (1.86, 2.64) | 5E-09 | 1 | 0 | 0 |
| PDZK1 | 118/118 | 2.96 (1.89, 4.65) | 6E-06 | 31/42,645 | 1.85 (1.57, 2.18) | 4E-07 | 1 | 0 | N/A |
| OSCAR | 118/118 | 2.72 (1.79, 4.15) | 8E-06 | 35/49,594 | 2.54 (1.87, 3.44) | 3E-06 | 1 | 0 | N/A |
| IGFBPL1 | 118/118 | 3.23 (1.97, 5.30) | 8E-06 | 35/49,618 | 2.31 (1.90, 2.82) | 2E-08 | 1 | 0 | N/A |
| SDC1 | 118/118 | 2.91 (1.84, 4.59) | 1E-05 | 35/49,618 | 2.04 (1.75, 2.36) | 2E-09 | 1 | 0 | N/A |
| PKD1 | 118/118 | 2.40 (1.64, 3.49) | 1E-05 | 31/42,773 | 2.79 (2.17, 3.59) | 1E-07 | 1 | 0 | N/A |
| VWF | 118/118 | 2.84 (1.82, 4.45) | 1E-05 | 35/49,746 | 1.89 (1.36, 2.63) | 6E-04 | 0 | 0 | 1 |
| RBKS | 118/118 | 2.96 (1.91, 4.58) | 4E-06 | 34/49,247 | 1.64 (1.18, 2.27) | 5E-03 | 0 | 0 | N/A |
| TP53I3 | 118/118 | 3.51 (2.05, 5.99) | 1E-05 | 31/42,782 | 1.44 (1.02, 2.04) | 4E-02 | 0 | 0 | N/A |

PLCO: Conditional logistic regression models of each protein's association with HCC, conditioned on matching factors, and adjusted for age, smoking, alcohol consumption, coffee consumption, education, BMI, diabetes. Missing covariates were imputed using MICE.

UK Biobank: Cox regression models of each protein's association with HCC, with follow-up time as the underlying variable. Models adjusted for age, sex, race, alcohol consumption, coffee consumption, smoking, education, BMI, and diabetes. Missing covariates were imputed using MICE.

Proteins listed here are those Bonferroni significant in PLCO ($p < 1.25\times10^{-5}$) and directionally consistent and $p < 0.05$ in UK Biobank.

*Restricting analyses to cases diagnosed ≥5 years after blood collection did not materially attenuate risk estimates in either cohort, defined as a change in the odds ratio of > –0.05 for positive associations or < 0.05 for inverse associations.

†Associations were considered consistent if $p < 0.05$ and directionally concordant with the largest prospective proteomic study of HCC to date (1,305 proteins; 54 cases, 54 controls)[17].

Associations for all proteins are available in the **Supplementary Data**.

Abbreviations: CI=confidence interval, HCC=hepatocellular carcinoma, HR=hazard ratio, MICE=multiple imputation by chained equations, OR=odds ratio, PLCO=Prostate, Lung, Colon, Ovary Screening Trial, SD=standard deviation

**Supplementary Table 2: Beta coefficients derived from LASSO models in PLCO**

| Assay | Coefficient |
|---|---|
| ADGRG1 | 0.3173 |
| LGALS3BP | 0.1330 |
| THBS2 | 0.1171 |
| TYMP | 0.1114 |
| ADAMTSL2 | 0.0341 |
| TSPAN8 | 0.0045 |
| HPD | 0.0012 |
| IGFBP3 | -0.0642 |

Models were trained using 3-fold cross-validation repeated 5 times, and performance was assessed using Cohen's Kappa, with final models selected based on the optimal lambda values

Abbreviations: ADAMTSL2=ADAMTS-like protein 2, ADGRG1=adhesion G-protein coupled receptor G1, HPD=4-hydroxyphenylpyruvate dioxygenase, IGFBP3=insulin-like growth factor binding protein-3, LASSO=Least Absolute Shrinkage and Selection Operator, LGALS3BP=galectin-3-binding protein, PLCO= Prostate, Lung, Colorectal, and Ovarian Cancer Screening Trial, THBS2=thrombospondin-2, TSPAN8=tetraspanin-8, TYMP=thymidine phosphorylase

**Supplementary Table 3: Associations of individual components of the multiprotein score with hepatocellular carcinoma risk factors in the UK Biobank**

| | | ADGRG1 | | LGALS3BP | | THBS2 | | TYMP | | ADAMTSL2 | | TSPAN8 | | IGFBP3 | |
|---|---|---|---|---|---|---|---|---|---|---|---|---|---|---|---|
| Risk factor | N | β (95% CI) | P | β (95% CI) | P | β (95% CI) | P | β (95% CI) | P | β (95% CI) | P | β (95% CI) | P | β (95% CI) | P |
| Age, years | | | | | | | | | | | | | | | |
| <45 | 4,409 | 0 (ref) | | 0 (ref) | | 0 (ref) | | 0 (ref) | | 0 (ref) | | 0 (ref) | | 0 (ref) | |
| 45-49 | 5,426 | 0.05 (0.02, 0.09) | **4e-03** | 0.02 (-0.01, 0.06) | 0.22 | 0.03 (-0.01, 0.07) | 0.13 | 0.01 (-0.03, 0.05) | 0.70 | 0.06 (0.02, 0.09) | **2e-03** | 0.1 (0.06, 0.14) | **3e-07** | -0.06 (-0.1, -0.03) | **6e-04** |
| 50-54 | 6,223 | 0.15 (0.11, 0.18) | **2e-17** | 0.05 (0.01, 0.09) | **6e-03** | 0.01 (-0.03, 0.04) | 0.67 | 0.05 (0.01, 0.09) | **0.01** | 0.16 (0.13, 0.2) | **6e-20** | 0.25 (0.21, 0.29) | **1e-39** | -0.07 (-0.11, -0.04) | **8e-05** |
| 55-59 | 7,456 | 0.27 (0.23, 0.3) | **4e-58** | 0.04 (0.01, 0.08) | **0.01** | -0.02 (-0.05, 0.02) | 0.32 | 0.03 (-0.01, 0.06) | 0.14 | 0.24 (0.21, 0.28) | **5e-45** | 0.32 (0.28, 0.35) | **1e-65** | -0.08 (-0.12, -0.05) | **5e-06** |
| 60-64 | 10,009 | 0.46 (0.43, 0.49) | **2e-180** | 0.07 (0.04, 0.1) | **8e-05** | 0.01 (-0.03, 0.04) | 0.69 | 0.05 (0.01, 0.08) | **7e-03** | 0.31 (0.28, 0.34) | **5e-78** | 0.34 (0.3, 0.37) | **7e-81** | -0.18 (-0.22, -0.15) | **1e-26** |
| 65+ | 8,453 | 0.72 (0.69, 0.76) | **<1e-300** | 0.09 (0.06, 0.13) | **2e-07** | 0.05 (0.01, 0.08) | **7e-03** | 0.07 (0.04, 0.11) | **5e-05** | 0.36 (0.33, 0.4) | **6e-101** | 0.38 (0.34, 0.41) | **1e-94** | -0.32 (-0.36, -0.29) | **7e-73** |
| Sex | | | | | | | | | | | | | | | |
| Women | 22,479 | 0 (ref) | | 0 (ref) | | 0 (ref) | | 0 (ref) | | 0 (ref) | | 0 (ref) | | 0 (ref) | |
| Men | 19,497 | 0.22 (0.21, 0.24) | **2e-127** | -0.22 (-0.24, -0.2) | **3e-101** | -0.25 (-0.27, -0.23) | **2e-128** | 0.01 (-0.01, 0.03) | 0.36 | -0.15 (-0.17, -0.13) | **2e-54** | 0.39 (0.37, 0.41) | **8e-305** | -0.52 (-0.54, -0.5) | **<1e-300** |
| Race | | | | | | | | | | | | | | | |
| White | 39,171 | 0 (ref) | | 0 (ref) | | 0 (ref) | | 0 (ref) | | 0 (ref) | | 0 (ref) | | 0 (ref) | |
| Asian | 965 | 0.11 (0.06, 0.17) | **1e-04** | 0.38 (0.32, 0.45) | **2e-34** | -0.18 (-0.24, -0.12) | **2e-08** | 0.21 (0.15, 0.28) | **6e-11** | 0.12 (0.07, 0.18) | **3e-05** | -0.07 (-0.13, -0.01) | **0.03** | -0.23 (-0.3, -0.17) | **1e-13** |
| Black | 1,040 | -0.21 (-0.27, -0.16) | **4e-14** | 0.12 (0.06, 0.18) | **5e-05** | -0.37 (-0.43, -0.31) | **1e-33** | 0.11 (0.05, 0.17) | **5e-04** | -0.27 (-0.33, -0.22) | **5e-21** | 0.02 (-0.04, 0.08) | 0.46 | -0.6 (-0.66, -0.54) | **2e-88** |
| Mixed race | 276 | -0.1 (-0.2, 0.01) | 0.07 | 0.08 (-0.03, 0.2) | 0.15 | -0.06 (-0.18, 0.05) | 0.27 | 0.07 (-0.05, 0.18) | 0.25 | 0.02 (-0.09, 0.13) | 0.71 | 0.05 (-0.06, 0.17) | 0.38 | -0.17 (-0.28, -0.06) | **3e-03** |
| Other | 524 | 0.02 (-0.06, 0.09) | 0.66 | 0.22 (0.14, 0.31) | **1e-07** | -0.19 (-0.27, -0.11) | **7e-06** | 0.06 (-0.03, 0.14) | 0.17 | -0.06 (-0.13, 0.02) | 0.17 | 0.01 (-0.07, 0.09) | 0.80 | -0.27 (-0.36, -0.19) | **4e-11** |
| BMI, $kg/m^2$ | | | | | | | | | | | | | | | |
| <18.5 | 219 | 0.1 (-0.01, 0.22) | 0.08 | -0.2 (-0.32, -0.07) | **2e-03** | 0.56 (0.43, 0.69) | **8e-18** | -0.06 (-0.19, 0.07) | 0.35 | 0.27 (0.15, 0.39) | **1e-05** | 0.23 (0.11, 0.36) | **4e-04** | -0.14 (-0.27, -0.02) | **0.03** |
| 18.5-24.9 | 13,454 | 0 (ref) | | 0 (ref) | | 0 (ref) | | 0 (ref) | | 0 (ref) | | 0 (ref) | | 0 (ref) | |
| 25-29.9 | 18,048 | 0.13 (0.11, 0.15) | **1e-39** | 0.28 (0.26, 0.31) | **2e-147** | -0.09 (-0.11, -0.06) | **1e-14** | 0.04 (0.02, 0.06) | **3e-04** | 0.16 (0.14, 0.18) | **5e-52** | 0.01 (-0.01, 0.03) | 0.34 | 0.12 (0.1, 0.14) | **5e-27** |
| 30-34.9 | 7,292 | 0.44 (0.42, 0.47) | **6e-252** | 0.63 (0.6, 0.65) | **<1e-300** | 0.17 (0.14, 0.2) | **9e-34** | 0.12 (0.1, 0.15) | **6e-18** | 0.58 (0.55, 0.6) | **<1e-300** | 0.06 (0.03, 0.09) | **3e-05** | 0.11 (0.08, 0.14) | **5e-15** |
| 35-39.9 | 2,107 | 0.78 (0.74, 0.82) | **1e-308** | 0.91 (0.86, 0.95) | **<1e-300** | 0.52 (0.47, 0.56) | **2e-113** | 0.17 (0.12, 0.21) | **7e-13** | 1.03 (0.99, 1.07) | **<1e-300** | 0.07 (0.02, 0.11) | **3e-03** | -0.04 (-0.08, 0.01) | 0.11 |
| 40+ | 856 | 0.98 (0.92, 1.04) | **3e-211** | 0.99 (0.92, 1.06) | **8e-186** | 0.74 (0.67, 0.8) | **2e-101** | 0.24 (0.17, 0.31) | **7e-12** | 1.29 (1.23, 1.36) | **1e-308** | 0.09 (0.02, 0.15) | **0.01** | -0.17 (-0.24, -0.11) | **2e-07** |
| Education | | | | | | | | | | | | | | | |
| <College | 11,499 | 0 (ref) | | 0 (ref) | | 0 (ref) | | 0 (ref) | | 0 (ref) | | 0 (ref) | | 0 (ref) | |
| College+ | 30,477 | -0.03 (-0.05, 0) | **0.04** | -0.04 (-0.06, -0.01) | **2e-03** | -0.01 (-0.03, 0.01) | 0.53 | -0.03 (-0.05, 0) | 0.06 | -0.03 (-0.05, -0.01) | **0.01** | -0.02 (-0.04, 0) | 0.12 | 0 (-0.03, 0.03) | 0.79 |
| Alcohol (drinks/d) | | | | | | | | | | | | | | | |
| < 1 | 25,886 | 0 (ref) | | 0 (ref) | | 0 (ref) | | 0 (ref) | | 0 (ref) | | 0 (ref) | | 0 (ref) | |
| 1-2 | 8,974 | -0.06 (-0.08, -0.04) | **2e-07** | 0.02 (-0.01, 0.04) | 0.15 | -0.02 (-0.05, 0) | 0.07 | 0 (-0.03, 0.02) | 0.84 | -0.06 (-0.08, -0.03) | **1e-06** | 0.02 (-0.01, 0.04) | 0.18 | 0.08 (0.05, 0.1) | **6e-11** |
| >2 | 7,116 | -0.07 (-0.09, -0.04) | **9e-08** | 0.14 (0.11, 0.17) | **2e-24** | 0.06 (0.04, 0.09) | **6e-06** | 0.06 (0.03, 0.09) | **1e-05** | 0.13 (0.11, 0.16) | **5e-24** | 0.12 (0.09, 0.14) | **2e-16** | 0.12 (0.09, 0.14) | **4e-17** |
| Coffee (cups/d) | | | | | | | | | | | | | | | |
| 0 | 9,461 | 0 (ref) | | 0 (ref) | | 0 (ref) | | 0 (ref) | | 0 (ref) | | 0 (ref) | | 0 (ref) | |
| 1-2 | 19,390 | -0.01 (-0.04, 0.01) | 0.21 | -0.08 (-0.1, -0.06) | **6e-11** | -0.04 (-0.06, -0.01) | **3e-03** | -0.02 (-0.04, 0.01) | 0.16 | -0.07 (-0.1, -0.05) | **3e-10** | 0.01 (-0.01, 0.04) | 0.34 | 0.02 (-0.01, 0.04) | 0.14 |
| 3-4 | 8,523 | -0.04 (-0.07, -0.02) | **1e-03** | -0.13 (-0.16, -0.1) | **7e-19** | -0.1 (-0.13, -0.07) | **7e-11** | 0 (-0.04, 0.03) | 0.77 | -0.15 (-0.18, -0.12) | **4e-26** | 0 (-0.03, 0.03) | 0.88 | -0.03 (-0.06, 0) | 0.06 |
| 5+ | 4,602 | -0.08 (-0.12, -0.05) | **6e-07** | -0.15 (-0.19, -0.12) | **9e-17** | -0.15 (-0.18, -0.11) | **1e-15** | -0.02 (-0.05, 0.02) | 0.37 | -0.17 (-0.21, -0.14) | **1e-23** | -0.01 (-0.05, 0.03) | 0.55 | -0.07 (-0.11, -0.04) | **9e-05** |
| Tea (cups/d) | | | | | | | | | | | | | | | |
| 0 | 6,105 | 0 (ref) | | 0 (ref) | | 0 (ref) | | 0 (ref) | | 0 (ref) | | 0 (ref) | | 0 (ref) | |
| 1-2 | 10,866 | -0.01 (-0.04, 0.02) | 0.43 | -0.01 (-0.04, 0.02) | 0.44 | -0.04 (-0.07, -0.01) | 0.01 | -0.03 (-0.06, 0) | 0.05 | -0.04 (-0.07, -0.01) | **4e-03** | -0.01 (-0.04, 0.02) | 0.44 | 0.06 (0.03, 0.09) | **3e-04** |
| 3-4 | 12,328 | -0.02 (-0.05, 0.01) | 0.22 | 0 (-0.04, 0.03) | 0.74 | -0.06 (-0.09, -0.03) | **1e-04** | -0.02 (-0.05, 0.01) | 0.30 | -0.07 (-0.1, -0.04) | **5e-07** | -0.03 (-0.06, 0) | **0.04** | 0.06 (0.03, 0.09) | **3e-04** |
| 5+ | 12,677 | -0.04 (-0.07, -0.02) | **2e-03** | -0.03 (-0.06, 0) | 0.08 | -0.09 (-0.12, -0.06) | **2e-08** | -0.01 (-0.04, 0.02) | 0.58 | -0.1 (-0.14, -0.08) | **2e-12** | -0.03 (-0.06, 0) | 0.09 | 0.03 (0, 0.06) | **0.03** |

| | | | | | | | | | | | | | | | |
|---|---|---|---|---|---|---|---|---|---|---|---|---|---|---|---|
| Smoking status | | | | | | | | | | | | | | | |
| Never | 22,825 | 0 (ref) | | 0 (ref) | | 0 (ref) | | 0 (ref) | | 0 (ref) | | 0 (ref) | | 0 (ref) | |
| Previous | 14,609 | 0.05 (0.03, 0.06) | **1e-06** | 0.04 (0.02, 0.06) | **2e-05** | 0.02 (0, 0.04) | **0.04** | 0.02 (0, 0.04) | 0.06 | 0.04 (0.03, 0.06) | **4e-06** | 0.04 (0.02, 0.06) | **1e-04** | -0.03 (-0.04, 0) | **0.01** |
| Current, <15 cig/d | 2,747 | 0.03 (0, 0.07) | 0.07 | 0.05 (0.01, 0.08) | **0.02** | 0.14 (0.1, 0.18) | **1e-12** | 0.06 (0.03, 0.1) | **1e-03** | 0.23 (0.2, 0.27) | **1e-35** | 0.13 (0.09, 0.17) | **2e-11** | -0.1 (-0.14, -0.06) | **9e-08** |
| Current, 15+ cig/d | 1,771 | 0.04 (0, 0.09) | 0.05 | 0.17 (0.12, 0.22) | **1e-12** | 0.26 (0.21, 0.3) | **4e-26** | 0.19 (0.15, 0.24) | **3e-15** | 0.46 (0.41, 0.5) | **2e-88** | 0.16 (0.11, 0.2) | **2e-10** | -0.15 (-0.19, -0.1) | **6e-10** |
| Current unknown | 24 | 0.16 (-0.19, 0.5) | 0.38 | 0.21 (-0.17, 0.59) | 0.28 | 0.49 (0.11, 0.87) | **0.01** | 0.09 (-0.3, 0.48) | 0.66 | 0.78 (0.42, 1.14) | **2e-05** | 0.24 (-0.15, 0.62) | 0.23 | -0.12 (-0.5, 0.25) | 0.52 |
| Diabetes | | | | | | | | | | | | | | | |
| No | 39,654 | 0 (ref) | | 0 (ref) | | 0 (ref) | | 0 (ref) | | 0 (ref) | | 0 (ref) | | 0 (ref) | |
| Yes | 2,322 | 0.98 (0.95, 1.02) | **<1e-300** | 0.22 (0.18, 0.26) | **2e-25** | 0.49 (0.45, 0.53) | **2e-120** | 0.08 (0.04, 0.12) | **2e-04** | 0.67 (0.63, 0.71) | **1e-248** | -0.13 (-0.17, -0.09) | **8e-10** | -0.42 (-0.46, -0.38) | **2e-91** |
| Hepatitis B | | | | | | | | | | | | | | | |
| No | 41,954 | 0 (ref) | | 0 (ref) | | 0 (ref) | | 0 (ref) | | 0 (ref) | | 0 (ref) | | 0 (ref) | |
| Yes | 22 | 0.77 (0.4, 1.14) | **4e-05** | 0.3 (-0.1, 0.7) | 0.15 | -0.09 (-0.5, 0.31) | 0.65 | 0.26 (-0.15, 0.68) | 0.21 | 0.06 (-0.32, 0.44) | 0.74 | 0 (-0.41, 0.4) | 0.99 | -0.26 (-0.66, 0.13) | 0.19 |
| Hepatitis C | | | | | | | | | | | | | | | |
| No | 41,951 | 0 (ref) | | 0 (ref) | | 0 (ref) | | 0 (ref) | | 0 (ref) | | 0 (ref) | | 0 (ref) | |
| Yes | 25 | 2.04 (1.7, 2.39) | **2e-31** | 1.89 (1.51, 2.26) | **4e-23** | 0.56 (0.19, 0.94) | **3e-03** | 0.44 (0.06, 0.83) | **0.02** | 0.66 (0.31, 1.02) | **2e-04** | 0.49 (0.11, 0.87) | **0.01** | -1.45 (-1.82, -1.08) | **2e-14** |
| ALD | | | | | | | | | | | | | | | |
| No | 41,929 | 0 (ref) | | 0 (ref) | | 0 (ref) | | 0 (ref) | | 0 (ref) | | 0 (ref) | | 0 (ref) | |
| Yes | 47 | 1.04 (0.79, 1.3) | **2e-15** | 0.42 (0.14, 0.7) | **3e-03** | 1.18 (0.89, 1.46) | **3e-16** | -0.05 (-0.34, 0.24) | 0.73 | 1.04 (0.78, 1.31) | **2e-14** | 0.63 (0.35, 0.92) | **1e-05** | -1.31 (-1.59, -1.03) | **2e-20** |
| MASLD | | | | | | | | | | | | | | | |
| No | 41,931 | 0 (ref) | | 0 (ref) | | 0 (ref) | | 0 (ref) | | 0 (ref) | | 0 (ref) | | 0 (ref) | |
| Yes | 45 | 0.5 (0.24, 0.75) | **1e-04** | 0.43 (0.16, 0.71) | **2e-03** | 0.43 (0.15, 0.71) | **3e-03** | 0.1 (-0.18, 0.39) | 0.49 | 0.6 (0.33, 0.86) | **1e-05** | 0.49 (0.21, 0.77) | **7e-04** | -0.29 (-0.57, -0.02) | **0.04** |
| Cirrhosis, hepatic fibrosis and sclerosis | | | | | | | | | | | | | | | |
| No | 41,911 | 0 (ref) | | 0 (ref) | | 0 (ref) | | 0 (ref) | | 0 (ref) | | 0 (ref) | | 0 (ref) | |
| Yes | 65 | 0.92 (0.7, 1.14) | **2e-16** | 0.52 (0.28, 0.76) | **2e-05** | 1.27 (1.03, 1.51) | **5e-25** | 0.06 (-0.18, 0.31) | 0.62 | 0.99 (0.77, 1.22) | **9e-18** | 0.94 (0.7, 1.19) | **3e-14** | -0.67 (-0.9, -0.43) | **3e-08** |

Associations estimated using linear models based on variables at study baseline. All models were mutually adjusted for the other variables. Missing data were imputed using multiple imputation by chained equations.

Abbreviations: ADAMTSL2=ADAMTS-like protein 2, ADGRG1=adhesion G-protein coupled receptor G1, ALD=alcohol-associated liver disease, BMI=body mass index, CI=confidence interval, IGFBP3=insulin-like growth factor binding protein-3, LGALS3BP=galectin-3-binding protein, MASLD=metabolic dysfunction-Associated Steatotic Liver Disease, SD=standard deviation, THBS2=thrombospondin-2, TSPAN8=tetraspanin-8, TYMP=thymidine phosphorylase

**Supplementary Table 4: Sequential area under the curve for predicting hepatocellular carcinoma risk by study**

| Proteins | Predictive performance | |
|---|---|---|
| | PLCO, AUC (95% CI) | UK Biobank, C-index (95% CI) |
| ADGRG1 | 0.86 (0.81, 0.91) | 0.90 (0.81, 0.96) |
| ADGRG1, LGALS3BP | 0.88 (0.84, 0.92) | 0.91 (0.83, 0.97) |
| ADGRG1, LGALS3BP, THBS2 | 0.89 (0.85, 0.93) | 0.92 (0.86, 0.98) |
| ADGRG1, LGALS3BP, THBS2, TYMP | 0.90 (0.86, 0.94) | 0.92 (0.85, 0.97) |
| ADGRG1, LGALS3BP, THBS2, TYMP, IGFBP3 | 0.90 (0.86, 0.94) | 0.92 (0.85, 0.98) |
| ADGRG1, LGALS3BP, THBS2, TYMP, IGFBP3, ADAMTSL2 | 0.90 (0.86, 0.94) | 0.92 (0.85, 0.98) |
| ADGRG1, LGALS3BP, THBS2, TYMP, IGFBP3, ADAMTSL2, TSPAN8 | 0.90 (0.86, 0.94) | 0.92 (0.85, 0.98) |
| ADGRG1, LGALS3BP, THBS2, TYMP, IGFBP3, ADAMTSL2, TSPAN8, HPD | 0.90 (0.86, 0.94) | N/A |

Progressive improvement in predictive performance as proteins are sequentially added to the model. Proteins were ordered based on their absolute beta weights derived from LASSO models derived in PLCO. Predictive performance based on the AUC in PLCO and the C-index in UK Biobank.

Abbreviations: ADAMTSL2=ADAMTS-like protein 2, ADGRG1=adhesion G-protein coupled receptor G1, AUC=area under the curve, CI=confidence interval, HPD=4-hydroxyphenylpyruvate dioxygenase, IGFBP3=insulin-like growth factor binding protein-3, LASSO=Least Absolute Shrinkage and Selection Operator, LGALS3BP=galectin-3-binding protein, PLCO= Prostate, Lung, Colorectal, and Ovarian Cancer Screening Trial, THBS2=thrombospondin-2, TSPAN8=tetraspanin-8, TYMP=thymidine phosphorylase

**Supplementary Table 5: Associations of multiprotein score and alpha-fetoprotein with subsequent diagnosis of liver-related diseases and mortality in the UK Biobank**

| Condition | Cases/total | Multiprotein score | | Alpha fetoprotein | |
|---|---|---|---|---|---|
| | | HR per 1 SD (95% CI) | P-Value | HR per 1 SD (95% CI) | P-Value |
| All-cause mortality | 5,127/41,982 | 1.24 (1.21, 1.27) | $2 \times 10^{-73}$ | 1.00 (0.97, 1.03) | 0.88 |
| Viral and non-viral liver conditions | 1,221/41,595 | 1.57 (1.51, 1.64) | $1 \times 10^{-82}$ | 1.09 (1.03, 1.16) | 0.004 |
| Non-viral liver conditions | 1,202/41,744 | 1.58 (1.51, 1.65) | $9 \times 10^{-85}$ | 1.09 (1.03, 1.16) | 0.003 |
| Alcohol-associated liver disease | 129/41,933 | 2.30 (2.11, 2.52) | $4 \times 10^{-36}$ | 1.62 (1.36, 1.93) | $3 \times 10^{-7}$ |
| MASLD | 584/41,936 | 1.58 (1.49, 1.67) | $9 \times 10^{-49}$ | 1.04 (0.95, 1.13) | 0.37 |
| Liver conditions excluding cirrhosis* | 1,167/41,819 | 1.57 (1.5, 1.64) | $1 \times 10^{-80}$ | 1.08 (1.02, 1.15) | 0.008 |
| Cirrhosis, hepatic fibrosis and sclerosis | 211/41,912 | 2.24 (2.08, 2.41) | $7 \times 10^{-54}$ | 1.47 (1.28, 1.69) | $1.4 \times 10^{-7}$ |
| Viral hepatitis | 49/41,851 | 2.02 (1.69, 2.42) | $2 \times 10^{-9}$ | 1.22 (0.91, 1.65) | 0.18 |
| Hepatitis B | 30/41,959 | 1.85 (1.45, 2.36) | $7 \times 10^{-5}$ | 0.89 (0.59, 1.34) | 0.54 |
| Hepatitis C | 21/41,955 | 2.47 (1.88, 3.24) | $8 \times 10^{-5}$ | 1.83 (1.13, 2.98) | 0.02 |

*First incident liver disease diagnosis was not cirrhosis, but participants may have developed cirrhosis later

Cases identified from national hospital admission and death records (anywhere on record). Prevalent cases were excluded from each respective analysis (based on hospital records prior to baseline and responses to the baseline interview). Associations were estimated using Cox proportional hazard models with time as the underlying variable, adjusted for age at blood collection (continuous) sex, race (White, individuals from all other racial groups), alcohol consumption (≤1, >1 drinks/day), coffee (continuous), smoking (never, previous, current), education (no college or equivalent professional qualifications, college graduate+), BMI (<25, 25-<30, 30+ kg/m$^2$), and diabetes (no, yes).

Abbreviations: BMI=body mass index, CI=confidence interval, HR=hazard ratio, MASLD=metabolic dysfunction-associated steatotic liver disease, SD=standard deviation

**Supplementary Table 6: Identification of at-risk groups for liver cancer**

| Diagnosis | Classification | | | N | |
|---|---|---|---|---|---|
| | ICD-10 | ICD-9 | Self-report codes | Prevalent | Incident |
| Liver cancer | C22 | 155 | - | - | 77 |
| Viral and non-viral liver conditions | K70-K77<br>B16<br>B17.0-B17.1<br>B18.0- B18.2<br>B18.8- B18.9<br>B19.1-B19.2 | 570-573<br>070.2-070.3<br>070.41<br>070.44<br>070.51<br>070.54<br>070.7 | 1604<br>1155<br>1156<br>1157<br>1158<br>1506<br>1580<br>1579 | 487 | 1,475 |
| Non-viral liver conditions | K70-K77 | 570-573 | 1604<br>1157<br>1158<br>1506 | 309 | 1,452 |
| Alcoholic liver disease | K70: Alcoholic liver disease (alcoholic fatty liver, alcoholic hepatitis, alcoholic fibrosis and sclerosis of liver, alcoholic cirrhosis of liver, alcoholic hepatic failure, alcoholic liver disease, unspecified) | 571.0: Alcoholic fatty liver<br>571.1: Acute alcoholic hepatitis<br>571.2: Alcoholic cirrhosis of liver<br>571.3: Alcoholic liver damage, unspecified | 1604 | 57 | 156 |
| MAFLD (including MASH) | K76.0: Fatty (change of) liver, not elsewhere classified<br>K75.8: Other specified inflammatory liver diseases | 571.8: Other chronic nonalcoholic liver disease<br>573.8: Other specified disorders of liver | - | 66 | 725 |
| Cirrhosis, hepatic fibrosis and sclerosis | K70.3: Alcoholic cirrhosis of liver<br>K74.0: Hepatic fibrosis<br>K74.1: Hepatic sclerosis<br>K74.2: Hepatic fibrosis with hepatic sclerosis<br>K74.3: Primary biliary cirrhosis<br>K74.4: Secondary biliary cirrhosis<br>K74.5: Biliary cirrhosis, unspecified K74.6: Other and unspecified cirrhosis of liver | 571.2: Alcoholic cirrhosis of liver<br>571.5: Cirrhosis of liver without mention of alcohol<br>571.6: Biliary cirrhosis | 1158<br>1506 | 91 | 253 |
| Non-viral diseases of the liver excluding advanced disease* | K70.0-K70.2<br>K70.4-K70.9<br>K71-K73.9<br>K74.7-K77 | 570-571.1<br>571.3-571.4<br>571.7-573 | 1604<br>1157 | 210 | 1,409 |
| Hepatitis B or C | B16: Acute hepatitis B | 070.2: Viral hepatitis B with hepatic coma | 1156 | 159 | 61 |

| | | | | | |
|---|---|---|---|---|---|
| | B17.0: Acute delta-(super) infection of hepatitis B carrier<br>B17.1: Acute hepatitis C<br>B18.0: Chronic viral hepatitis B with delta-agent<br>B18.1: Chronic viral hepatitis B without delta-agent<br>B18.2: Chronic viral hepatitis C<br>B18.8: Other chronic viral hepatitis<br>B18.9: Chronic viral hepatitis, unspecified<br>B19.1: Unspecified viral hepatitis B<br>B19.2: Unspecified viral hepatitis C | 070.3: Viral hepatitis B without mention of hepatic coma<br>070.41: Acute hepatitis C with hepatic coma<br>070.44: Chronic hepatitis C with hepatic coma<br>070.51: Acute hepatitis C without mention of hepatic coma<br>070.54: Chronic hepatitis C without mention of hepatic coma<br>070.7: Unspecified viral hepatitis C | 1580<br>1579 | | |
| Hepatitis B | B16: Acute hepatitis B<br>B17.0: Acute delta-(super) infection of hepatitis B carrier<br>B18.0: Chronic viral hepatitis B with delta-agent<br>B18.1: Chronic viral hepatitis B without delta-agent<br>B19.1: Unspecified viral hepatitis B | 070.2: Viral hepatitis B with hepatic coma<br>070.3: Viral hepatitis B without mention of hepatic coma | 1579 | 29 | 40 |
| Hepatitis C | B17.1: Acute hepatitis C<br>B18.2: Chronic viral hepatitis C<br>B19.2: Unspecified viral hepatitis C | 070.41: Acute hepatitis C with hepatic coma<br>070.44: Chronic hepatitis C with hepatic coma<br>070.51: Acute hepatitis C without mention of hepatic coma<br>070.54: Chronic hepatitis C without mention of hepatic coma<br>070.7: Unspecified viral hepatitis C | 1580 | 28 | 23 |

ICD-9 and ICD-10 codes were used to identify conditions from hospital admission records and death records (anywhere on record). Self-report codes are based on the interview assessment, coded by UK Biobank (variable 20002_0), more information is available from: https://biobank.ndph.ox.ac.uk/showcase/field.cgi?id=20002

*First incident liver disease diagnosis was not cirrhosis or related condition, but participants may have developed cirrhosis later

Abbreviations: MASLD=metabolic dysfunction-associated steatotic liver disease, MASH=metabolic dysfunction–associated steatohepatitis

**Supplementary Table 7: Annual risks of hepatocellular carcinoma and liver cancer over a 7-year period**

| Condition | Hepatocellular carcinoma, % | Liver cancer, % |
|---|---|---|
| Viral and non-viral liver conditions | 0.21 | 0.27 |
| Non-viral liver conditions | 0.26 | 0.33 |
| Non-viral liver diseases excluding cirrhosis, fibrosis and sclerosis* | 0.24 | 0.3 |
| Cirrhosis, hepatic fibrosis and sclerosis | 0.97 | 1.08 |
| Hepatitis B or C | 0.35 | 0.43 |

*First incident liver disease diagnosis was not cirrhosis or related condition, but participants may have developed cirrhosis later

Analysis restricted to liver-related conditions with at least 5 HCC cases. The cumulative risk of cancer was estimated using a Cox proportional hazards regression model with time as the underlying variable. Follow-up time was calculated from baseline blood collection for participants with prevalent disease (hospital admission records or self-report at baseline) or from first diagnosis if the condition occurred after study baseline (hospital admission records).

The baseline cumulative hazard was determined at year 7. Individual predicted risks were then derived using the survival function and linear predictor from the Cox model. Finally, the population-level cumulative risk was calculated as the average of these individual predictions and divided by 7 to calculate annual risk.

**Supplementary Table 8: Associations of the multiprotein score with liver cancer and HCC risk in the overall and at-risk populations in the UK Biobank**

| Study population | Site | N cases/ total | Protein risk score | | | Alpha-fetoprotein | | |
|---|---|---|---|---|---|---|---|---|
| | | | HR per 1 SD (95% CI) | C-index (95% CI) | P-Value | HR per 1 SD (95% CI) | C-index (95% CI) | P-Value |
| Overall population | Liver cancer | 60/41,978 | 2.22 (2.01, 2.45) | 0.76 (0.68, 0.84) | $4.1 \times 10^{-57}$ | 1.60 (1.26, 2.02) | 0.60 (0.54, 0.67) | 0.0001 |
| | HCC | 28/41,982 | 2.65 (2.35, 2.99) | 0.92 (0.84, 0.97) | $2.4 \times 10^{-56}$ | 1.95 (1.40, 2.71) | 0.66 (0.55, 0.76) | 0.00007 |
| Viral and non-viral liver conditions | Liver cancer | 23/1,562 | 1.82 (1.58, 2.09) | 0.83 (0.69, 0.95) | $6 \times 10^{-17}$ | 1.94 (1.34, 2.82) | 0.63 (0.50, 0.76) | 0.0005 |
| | HCC | 16/1,546 | 2.00 (1.69, 2.36) | 0.94 (0.91, 0.97) | $3 \times 10^{-16}$ | 2.36 (1.53, 3.64) | 0.65 (0.50, 0.82) | 0.0001 |
| Non-viral liver conditions | Liver cancer | 22/1,395 | 1.80 (1.56, 2.08) | 0.83 (0.68, 0.95) | $1 \times 10^{-15}$ | 1.97 (1.35, 2.87) | 0.63 (0.50, 0.78) | 0.0004 |
| | HCC | 15/1,378 | 2.00 (1.68, 2.38) | 0.94 (0.91, 0.97) | $3 \times 10^{-15}$ | 2.45 (1.56, 3.82) | 0.66 (0.49, 0.85) | 0.00009 |
| Cirrhosis, hepatic fibrosis and sclerosis | Liver cancer | 14/268 | 1.58 (1.31, 1.90) | 0.83 (0.72, 0.93) | $2 \times 10^{-6}$ | 2.00 (1.26, 3.17) | 0.67 (0.52, 0.81) | 0.003 |
| | HCC | 11/265 | 1.69 (1.36, 2.08) | 0.88 (0.81, 0.95) | $1 \times 10^{-6}$ | 2.26 (1.36, 3.76) | 0.67 (0.49, 0.87) | 0.002 |
| Non-viral liver conditions excluding cirrhosis* | Liver cancer | 18/1,288 | 1.77 (1.51, 2.08) | 0.82 (0.64, 0.95) | $3 \times 10^{-12}$ | 2.21 (1.47, 3.30) | 0.65 (0.49, 0.83) | 0.0001 |
| | HCC | 13/1,271 | 1.95 (1.62, 2.35) | 0.94 (0.90, 0.97) | $2 \times 10^{-12}$ | 2.67 (1.68, 4.25) | 0.69 (0.50, 0.89) | 0.00003 |
| Hepatitis B or C | Liver cancer | 6/177 | 1.77 (1.39, 2.25) | 0.94 (0.88, 0.99) | $4 \times 10^{-6}$ | 2.39 (1.39, 4.10) | 0.71 (0.48, 0.97) | 0.002 |
| | HCC | 5/177 | 1.74 (1.34, 2.26) | 0.93 (0.86, 1.00) | $3 \times 10^{-5}$ | 2.63 (1.46, 4.72) | 0.71 (0.46, 1.00) | 0.001 |

*First incident liver disease diagnosis was not cirrhosis, hepatitis fibrosis or sclerosis, but participants may have developed these conditions later

Restricted to liver-related conditions with at least 5 HCC cases and participants with complete protein score and alpha-fetoprotein measures. Associations were estimated using Cox proportional hazards models with follow-up time as the underlying variable. For populations with prevalent disease, entry time was defined as the date of baseline blood collection. For those diagnosed with a condition after study baseline, entry time was the date of first recorded diagnosis. Participants with cancer record prior to high-risk disease diagnosis were excluded.

Abbreviations: C-index=concordance index, HCC=hepatocellular carcinoma, HR=hazard ratio, SD=standard deviation

**Supplementary Table 9: Optimal thresholds, sensitivity, specificity and cumulative risk of the protein risk score in at-risk populations in the UK Biobank**

| Study population | Site | High-risk cutpoint (SD) | N cases above cutpoint/ total cases | N non-cases above/ total non-cases | Sensitivity | Specificity | Cumulative risk in participants with each condition | |
|---|---|---|---|---|---|---|---|---|
| | | | | | | | High-risk, % | Low-risk, % |
| Viral and non-viral liver conditions | Liver cancer | 2.10 | 20/23 | 288/1,539 | 0.716 | 0.813 | 7.31 | 0.71 |
| | HCC | 2.44 | 16/16 | 251/1,530 | 0.77 | 0.837 | 6.93 | 0.43 |
| Non-viral liver conditions | Liver cancer | 2.21 | 19/22 | 252/1,373 | 0.709 | 0.817 | 7.98 | 0.79 |
| | HCC | 2.55 | 14/15 | 220/1,363 | 0.773 | 0.84 | 7.51 | 0.45 |
| Cirrhosis, hepatic fibrosis and sclerosis | Liver cancer | 3.30 | 10/14 | 68/254 | 0.693 | 0.742 | 18.01 | 3.27 |
| | HCC | 3.51 | 9/11 | 64/254 | 0.729 | 0.757 | 17.56 | 2.48 |
| Non-viral liver conditions excluding cirrhosis* | Liver cancer | 2.10 | 16/18 | 241/1,270 | 0.701 | 0.811 | 7.01 | 0.75 |
| | HCC | 2.44 | 13/13 | 211/1,258 | 0.762 | 0.833 | 6.52 | 0.44 |
| Hepatitis B or C | Liver cancer | 2.73 | 4/6 | 23/171 | 0.714 | 0.866 | 15.20 | 1.10 |
| | HCC | 2.73 | 3/5 | 24/172 | 0.707 | 0.864 | 13.43 | 1.00 |

*First incident liver disease diagnosis was not cirrhosis, hepatitis fibrosis or sclerosis, but participants may have developed these conditions later

Analysis restricted to liver-related conditions with at least 5 HCC cases. Associations were estimated using Cox proportional hazard models with time as the underlying variable. Follow-up time was calculated from baseline blood collection for participants with prevalent disease or from date at first diagnosis if the condition occurred after study baseline, based on hospital admission records.

Sensitivity, specificity, and cumulative risk were evaluated up to 8 years follow-up time. Score metrics by year are available from **Supplementary Table 12** and details of calculations are available from the **Supplementary Methods**.

Cut-points derived using Youden's Index (maximum sensitivity + specificity – 1) at 3 years of follow-up.
High risk(t) = Cumulative risk for participants with a protein risk score ≥ cutpoint (PPV(t))
Low risk(t) = Cumulative risk for participants with a protein risk score < cutpoint (cNPV(t))
Sensitivity(t)= Proportion of participants ≥ cutpoint who developed the outcome, relative to the mean risk in the population
Specificity(t)= Proportion of participants < cutpoint who did not develop the outcome, relative to the mean risk in the population.

Abbreviations: CI=confidence interval, HCC=hepatocellular carcinoma, SD=standard deviation

**Supplementary Table 10: Optimal thresholds, sensitivity, specificity and cumulative risk of the protein risk score in at-risk populations with prevalent conditions at study baseline in the UK Biobank**

| Study population | Site | High-risk cutpoint (SD) | N cases above cutpoint/ total cases | N non-cases above/ total non-cases | Sensitivity | Specificity | Cumulative risk in participants with each condition | |
|---|---|---|---|---|---|---|---|---|
| | | | | | | | High-risk, % | Low-risk, % |
| Viral and non-viral liver conditions | Liver cancer | 2.32 | 12/13 | 71/374 | 0.695 | 0.8 | 9.43 | 1.13 |
| | HCC | 2.44 | 10/10 | 70/377 | 0.719 | 0.807 | 9.16 | 0.93 |
| Non-viral liver conditions | Liver cancer | 2.55 | 7/10 | 46/228 | 0.663 | 0.794 | 10.78 | 1.57 |
| | HCC | 2.66 | 6/7 | 44/231 | 0.697 | 0.806 | 10.32 | 1.19 |
| Cirrhosis, hepatic fibrosis and sclerosis | Liver cancer | 2.96 | 4/7 | 14/63 | 0.695 | 0.792 | 27.05 | 4.10 |
| | HCC | 3.38 | 3/5 | 13/65 | 0.73 | 0.814 | 25.20 | 2.77 |
| Hepatitis B or C | Liver cancer | 2.51 | 5/5 | 17/126 | 0.697 | 0.851 | 14.68 | 1.29 |
| | HCC | 2.51 | 4/4 | 18/127 | 0.68 | 0.848 | 12.37 | 1.17 |

Associations were estimated using Cox proportional hazard models with time as the underlying variable. Follow-up time was calculated from baseline blood collection for participants with prevalent disease only.

Sensitivity, specificity, and cumulative risk were evaluated up to 8 years follow-up time. Details of calculations are available from the **Supplementary Methods**.

Cut-points derived using Youden's Index (maximum sensitivity + specificity – 1) at 3 years of follow-up.
High risk(t) = Cumulative risk for participants with a protein risk score ≥ cutpoint (PPV(t))
Low risk(t) = Cumulative risk for participants with a protein risk score < cutpoint (cNPV(t))
Sensitivity(t)= Proportion of participants ≥ cutpoint who developed the outcome, relative to the mean risk in the population
Specificity(t)= Proportion of participants < cutpoint who did not develop the outcome, relative to the mean risk in the population.

Abbreviations: CI=confidence interval, HCC=hepatocellular carcinoma, SD=standard deviation

**Supplementary Table 11: Associations of alpha-fetoprotein with liver cancer and HCC risk in the overall and at-risk populations in the UK Biobank**

| Study population | Site | High risk cutpoint (SD) | N cases above cutpoint/ total | N non-cases above cutpoint/ total | Sensitivity | Specificity | Cumulative risk in participants with each condition | |
|---|---|---|---|---|---|---|---|---|
| | | | | | | | High-risk, % | Low-risk, % |
| Viral and non-viral liver conditions | Liver cancer | 0.45 | 13/23 | 504/1,539 | 0.599 | 0.675 | 3.76 | 1.25 |
| | HCC | 0.53 | 7/16 | 460/1,530 | 0.651 | 0.704 | 3.60 | 0.84 |
| Non-viral liver conditions | Liver cancer | 0.45 | 13/22 | 451/1,373 | 0.608 | 0.674 | 4.16 | 1.34 |
| | HCC | 0.61 | 7/15 | 364/1,363 | 0.635 | 0.737 | 4.26 | 0.90 |
| Cirrhosis, hepatic fibrosis and sclerosis | Liver cancer | 0.62 | 6/14 | 84/254 | 0.596 | 0.685 | 13.28 | 4.55 |
| | HCC | 0.69 | 5/11 | 75/254 | 0.609 | 0.72 | 13.26 | 3.68 |
| Non-viral liver conditions excluding cirrhosis* | Liver cancer | 0.53 | 9/18 | 380/1,270 | 0.627 | 0.705 | 4.34 | 1.12 |
| | HCC | 0.61 | 6/13 | 335/1,258 | 0.675 | 0.739 | 4.19 | 0.74 |
| Hepatitis B or C | Liver cancer | 0.75 | 3/6 | 39/171 | 0.636 | 0.778 | 9.94 | 1.77 |
| | HCC | 0.82 | 3/5 | 38/172 | 0.679 | 0.783 | 9.29 | 1.33 |

Analysis restricted to liver-related conditions with at least 5 HCC cases. Associations were estimated using Cox proportional hazard models with time as the underlying variable. Follow-up time was calculated from baseline blood collection for participants with prevalent disease or from date at first diagnosis if the condition occurred after study baseline, based on hospital admission records.

Sensitivity, specificity, and cumulative risk were evaluated up to 8 years follow-up time. Score metrics by year are available from **Supplementary Table 13** and details of calculations are available from the **Supplementary Methods**.

Cut-points derived using Youden's Index (maximum sensitivity + specificity – 1) at 3 years of follow-up
High risk(t) = Cumulative risk for participants with a protein risk score ≥ cutpoint (PPV(t))
Low risk(t) = Cumulative risk for participants with a protein risk score < cutpoint (cNPV(t))
Sensitivity(t)= Proportion of participants ≥ cutpoint who developed the outcome, relative to the mean risk in the populationSpecificity(t)= Proportion of participants < cutpoint who did not develop the outcome, relative to the mean risk in the population.

Abbreviations: CI=confidence interval, HCC=hepatocellular carcinoma, SD=standard deviation

**Supplementary Table 12: Sensitivity, specificity and cumulative risk over time for the protein risk score in at-risk populations in the UK Biobank**

| Condition | Cancer | High risk cut-point (SD) | Follow-up, yrs | Sensitivity | Specificity | Cumulative risk in participants with each condition | |
|---|---|---|---|---|---|---|---|
| | | | | | | High-risk, % | Low-risk, % |
| Viral and non-viral liver conditions | Liver cancer | 2.1 | 1 | 0.731 | 0.806 | 2.03 | 0.184 |
| | | | 2 | 0.73 | 0.806 | 2.35 | 0.214 |
| | | | 3 | 0.728 | 0.807 | 3.09 | 0.284 |
| | | | 4 | 0.725 | 0.808 | 3.894 | 0.362 |
| | | | 5 | 0.72 | 0.811 | 5.666 | 0.54 |
| | | | 6 | 0.719 | 0.812 | 6.156 | 0.59 |
| | | | 7 | 0.718 | 0.813 | 6.708 | 0.648 |
| | | | 8 | 0.716 | 0.813 | 7.313 | 0.712 |
| | | | 9 | 0.714 | 0.814 | 7.982 | 0.784 |
| | | | 10 | 0.712 | 0.816 | 8.754 | 0.868 |
| | HCC | 2.4 | 1 | 0.788 | 0.829 | 1.255 | 0.07 |
| | | | 2 | 0.787 | 0.83 | 1.645 | 0.093 |
| | | | 3 | 0.784 | 0.831 | 2.533 | 0.146 |
| | | | 4 | 0.781 | 0.832 | 3.482 | 0.204 |
| | | | 5 | 0.775 | 0.834 | 5.052 | 0.306 |
| | | | 6 | 0.774 | 0.835 | 5.627 | 0.344 |
| | | | 7 | 0.772 | 0.836 | 6.259 | 0.387 |
| | | | 8 | 0.77 | 0.837 | 6.932 | 0.433 |
| Non-viral liver conditions | Liver cancer | 2.2 | 1 | 0.723 | 0.809 | 2.511 | 0.231 |
| | | | 2 | 0.723 | 0.809 | 2.511 | 0.231 |
| | | | 3 | 0.721 | 0.811 | 3.402 | 0.317 |
| | | | 4 | 0.718 | 0.812 | 4.384 | 0.414 |
| | | | 5 | 0.714 | 0.814 | 6.012 | 0.58 |
| | | | 6 | 0.711 | 0.816 | 7.26 | 0.712 |
| | | | 7 | 0.709 | 0.817 | 7.98 | 0.789 |
| | | | 8 | 0.709 | 0.817 | 7.98 | 0.789 |
| | | | 9 | 0.707 | 0.818 | 8.837 | 0.883 |
| | | | 10 | 0.704 | 0.82 | 9.844 | 0.996 |
| | HCC | 2.6 | 1 | 0.792 | 0.832 | 1.705 | 0.092 |
| | | | 2 | 0.792 | 0.832 | 1.705 | 0.092 |
| | | | 3 | 0.788 | 0.834 | 2.793 | 0.154 |
| | | | 4 | 0.784 | 0.836 | 3.964 | 0.223 |
| | | | 5 | 0.78 | 0.837 | 5.245 | 0.302 |
| | | | 6 | 0.776 | 0.839 | 6.682 | 0.395 |
| | | | 7 | 0.773 | 0.84 | 7.507 | 0.45 |
| | | | 8 | 0.773 | 0.84 | 7.507 | 0.45 |
| Cirrhosis, hepatic fibrosis and sclerosis | Liver cancer | 3.3 | 1 | 0.716 | 0.716 | 3.945 | 0.643 |
| | | | 2 | 0.716 | 0.716 | 3.945 | 0.643 |
| | | | 3 | 0.708 | 0.724 | 8.582 | 1.45 |
| | | | 4 | 0.703 | 0.73 | 11.943 | 2.071 |
| | | | 5 | 0.703 | 0.73 | 11.943 | 2.071 |
| | | | 6 | 0.693 | 0.742 | 18.012 | 3.274 |
| | | | 7 | 0.693 | 0.742 | 18.012 | 3.274 |
| | | | 8 | 0.693 | 0.742 | 18.012 | 3.274 |
| | | | 9 | 0.689 | 0.747 | 20.703 | 3.844 |
| | | | 10 | 0.689 | 0.747 | 20.703 | 3.844 |
| | HCC | 3.5 | 1 | 0.756 | 0.728 | 2.133 | 0.261 |

| | | | | | | | |
|---|---|---|---|---|---|---|---|
| | | | 2 | 0.756 | 0.728 | 2.133 | 0.261 |
| | | | 3 | 0.747 | 0.738 | 7.314 | 0.941 |
| | | | 4 | 0.741 | 0.744 | 11.016 | 1.467 |
| | | | 5 | 0.741 | 0.744 | 11.016 | 1.467 |
| | | | 6 | 0.729 | 0.757 | 17.558 | 2.484 |
| | | | 7 | 0.729 | 0.757 | 17.558 | 2.484 |
| | | | 8 | 0.729 | 0.757 | 17.558 | 2.484 |
| Non-viral liver conditions excluding cirrhosis* | Liver cancer | 2.1 | 1 | 0.713 | 0.804 | 2.276 | 0.229 |
| | | | 2 | 0.713 | 0.804 | 2.276 | 0.229 |
| | | | 3 | 0.712 | 0.804 | 2.754 | 0.278 |
| | | | 4 | 0.709 | 0.806 | 3.812 | 0.391 |
| | | | 5 | 0.703 | 0.81 | 6.224 | 0.657 |
| | | | 6 | 0.703 | 0.81 | 6.224 | 0.657 |
| | | | 7 | 0.701 | 0.811 | 7.007 | 0.746 |
| | | | 8 | 0.701 | 0.811 | 7.007 | 0.746 |
| | | | 9 | 0.701 | 0.811 | 7.007 | 0.746 |
| | | | 10 | 0.698 | 0.812 | 8.225 | 0.889 |
| | HCC | 2.4 | 1 | 0.777 | 0.826 | 1.745 | 0.107 |
| | | | 2 | 0.777 | 0.826 | 1.745 | 0.107 |
| | | | 3 | 0.775 | 0.827 | 2.315 | 0.143 |
| | | | 4 | 0.771 | 0.829 | 3.561 | 0.226 |
| | | | 5 | 0.765 | 0.832 | 5.649 | 0.372 |
| | | | 6 | 0.765 | 0.832 | 5.649 | 0.372 |
| | | | 7 | 0.762 | 0.833 | 6.52 | 0.436 |
| | | | 8 | 0.762 | 0.833 | 6.52 | 0.436 |
| Hepatitis B or C | Liver cancer | 2.7 | 1 | 0.751 | 0.854 | 5.392 | 0.322 |
| | | | 2 | 0.742 | 0.857 | 7.842 | 0.491 |
| | | | 3 | 0.742 | 0.857 | 7.842 | 0.491 |
| | | | 4 | 0.733 | 0.86 | 10.201 | 0.668 |
| | | | 5 | 0.724 | 0.863 | 12.58 | 0.862 |
| | | | 6 | 0.724 | 0.863 | 12.58 | 0.862 |
| | | | 7 | 0.724 | 0.863 | 12.58 | 0.862 |
| | | | 8 | 0.714 | 0.866 | 15.204 | 1.096 |
| | | | 9 | 0.714 | 0.866 | 15.204 | 1.096 |
| | | | 10 | 0.714 | 0.866 | 15.204 | 1.096 |
| | HCC | 2.7 | 1 | 0.744 | 0.851 | 2.861 | 0.177 |
| | | | 2 | 0.735 | 0.854 | 5.51 | 0.357 |
| | | | 3 | 0.735 | 0.854 | 5.51 | 0.357 |
| | | | 4 | 0.727 | 0.857 | 8.059 | 0.546 |
| | | | 5 | 0.717 | 0.861 | 10.62 | 0.753 |
| | | | 6 | 0.717 | 0.861 | 10.62 | 0.753 |
| | | | 7 | 0.717 | 0.861 | 10.62 | 0.753 |
| | | | 8 | 0.707 | 0.864 | 13.431 | 1.003 |

*First incident liver disease diagnosis was not cirrhosis, hepatitis fibrosis or sclerosis, but participants may have developed these conditions later

Analysis restricted to liver-related conditions with at least 5 HCC cases. Associations were estimated using Cox proportional hazard models with time as the underlying variable. Follow-up time was calculated from baseline blood collection for participants with prevalent disease or from age at first diagnosis if the condition occurred after study baseline, based on hospital admission records.

Cut-points derived using Youden's Index (maximum sensitivity + specificity – 1) at 3 years of follow-up
High risk(t) = Cumulative risk for participants with a protein risk score ≥ cutpoint (PPV(t))
Low risk(t) = Cumulative risk for participants with a protein risk score < cutpoint (cNPV(t))
Sensitivity(t)= Proportion of participants ≥ cutpoint who developed the outcome, relative to the mean risk in the population
Specificity(t)= Proportion of participants < cutpoint who did not develop the outcome, relative to the mean risk in the population.

Abbreviations: CI=confidence interval, HCC=hepatocellular carcinoma

**Supplementary Table 13: Sensitivity, specificity and cumulative risk over time for alpha-fetoprotein in at-risk populations in the UK Biobank**

| Condition | Cancer | High risk cut-point (SD) | Follow-up, yrs | Sensitivity | Specificity | Cumulative risk in participants with each condition | |
|---|---|---|---|---|---|---|---|
| | | | | | | High-risk, % | Low-risk, % |
| Viral and non-viral liver conditions | Liver cancer | 0.45 | 1 | 0.602 | 0.671 | 1.025 | 0.335 |
| | | | 2 | 0.602 | 0.671 | 1.184 | 0.387 |
| | | | 3 | 0.602 | 0.671 | 1.552 | 0.508 |
| | | | 4 | 0.601 | 0.672 | 1.957 | 0.643 |
| | | | 5 | 0.6 | 0.673 | 2.877 | 0.949 |
| | | | 6 | 0.6 | 0.674 | 3.127 | 1.032 |
| | | | 7 | 0.599 | 0.674 | 3.419 | 1.13 |
| | | | 8 | 0.599 | 0.675 | 3.764 | 1.246 |
| | | | 9 | 0.599 | 0.675 | 4.144 | 1.375 |
| | | | 10 | 0.598 | 0.676 | 4.587 | 1.525 |
| | HCC | 0.53 | 1 | 0.656 | 0.699 | 0.611 | 0.139 |
| | | | 2 | 0.656 | 0.699 | 0.8 | 0.182 |
| | | | 3 | 0.655 | 0.7 | 1.233 | 0.281 |
| | | | 4 | 0.654 | 0.701 | 1.711 | 0.392 |
| | | | 5 | 0.653 | 0.702 | 2.543 | 0.585 |
| | | | 6 | 0.652 | 0.703 | 2.84 | 0.655 |
| | | | 7 | 0.652 | 0.703 | 3.188 | 0.737 |
| | | | 8 | 0.651 | 0.704 | 3.602 | 0.835 |
| Non-viral liver conditions | Liver cancer | 0.45 | 1 | 0.611 | 0.669 | 1.277 | 0.404 |
| | | | 2 | 0.611 | 0.669 | 1.277 | 0.404 |
| | | | 3 | 0.611 | 0.67 | 1.724 | 0.547 |
| | | | 4 | 0.61 | 0.671 | 2.229 | 0.709 |
| | | | 5 | 0.609 | 0.672 | 3.1 | 0.991 |
| | | | 6 | 0.608 | 0.673 | 3.779 | 1.212 |
| | | | 7 | 0.608 | 0.674 | 4.164 | 1.338 |
| | | | 8 | 0.608 | 0.674 | 4.164 | 1.338 |
| | | | 9 | 0.607 | 0.675 | 4.702 | 1.515 |
| | | | 10 | 0.606 | 0.676 | 5.357 | 1.732 |
| | HCC | 0.61 | 1 | 0.641 | 0.732 | 0.898 | 0.185 |
| | | | 2 | 0.641 | 0.732 | 0.898 | 0.185 |
| | | | 3 | 0.64 | 0.733 | 1.473 | 0.305 |
| | | | 4 | 0.639 | 0.734 | 2.123 | 0.441 |
| | | | 5 | 0.638 | 0.735 | 2.89 | 0.604 |
| | | | 6 | 0.636 | 0.737 | 3.764 | 0.792 |
| | | | 7 | 0.635 | 0.737 | 4.256 | 0.899 |
| | | | 8 | 0.635 | 0.737 | 4.256 | 0.899 |
| Cirrhosis, hepatic fibrosis and sclerosis | Liver cancer | 0.62 | 1 | 0.612 | 0.669 | 2.999 | 0.963 |
| | | | 2 | 0.612 | 0.669 | 2.999 | 0.963 |
| | | | 3 | 0.607 | 0.674 | 6.202 | 2.032 |
| | | | 4 | 0.603 | 0.677 | 8.503 | 2.825 |
| | | | 5 | 0.603 | 0.677 | 8.503 | 2.825 |
| | | | 6 | 0.596 | 0.685 | 13.278 | 4.545 |
| | | | 7 | 0.596 | 0.685 | 13.278 | 4.545 |
| | | | 8 | 0.596 | 0.685 | 13.278 | 4.545 |
| | | | 9 | 0.593 | 0.689 | 15.618 | 5.425 |
| | | | 10 | 0.593 | 0.689 | 15.618 | 5.425 |
| | HCC | 0.69 | 1 | 0.633 | 0.701 | 1.646 | 0.414 |
| | | | 2 | 0.633 | 0.701 | 1.646 | 0.414 |

| | | | | | | | |
|---|---|---|---|---|---|---|---|
| | | | 3 | 0.625 | 0.707 | 5.298 | 1.374 |
| | | | 4 | 0.62 | 0.711 | 7.902 | 2.096 |
| | | | 5 | 0.62 | 0.711 | 7.902 | 2.096 |
| | | | 6 | 0.609 | 0.72 | 13.26 | 3.68 |
| | | | 7 | 0.609 | 0.72 | 13.26 | 3.68 |
| | | | 8 | 0.609 | 0.72 | 13.26 | 3.68 |
| Non-viral liver conditions excluding cirrhosis* | Liver cancer | 0.53 | 1 | 0.631 | 0.7 | 1.369 | 0.346 |
| | | | 2 | 0.631 | 0.7 | 1.369 | 0.346 |
| | | | 3 | 0.631 | 0.701 | 1.648 | 0.417 |
| | | | 4 | 0.63 | 0.702 | 2.298 | 0.584 |
| | | | 5 | 0.627 | 0.704 | 3.834 | 0.985 |
| | | | 6 | 0.627 | 0.704 | 3.834 | 0.985 |
| | | | 7 | 0.627 | 0.705 | 4.342 | 1.119 |
| | | | 8 | 0.627 | 0.705 | 4.342 | 1.119 |
| | | | 9 | 0.627 | 0.705 | 4.342 | 1.119 |
| | | | 10 | 0.625 | 0.706 | 5.314 | 1.378 |
| | HCC | 0.61 | 1 | 0.681 | 0.733 | 1.037 | 0.178 |
| | | | 2 | 0.681 | 0.733 | 1.037 | 0.178 |
| | | | 3 | 0.681 | 0.734 | 1.371 | 0.236 |
| | | | 4 | 0.679 | 0.735 | 2.156 | 0.374 |
| | | | 5 | 0.676 | 0.738 | 3.576 | 0.628 |
| | | | 6 | 0.676 | 0.738 | 3.576 | 0.628 |
| | | | 7 | 0.675 | 0.739 | 4.185 | 0.74 |
| | | | 8 | 0.675 | 0.739 | 4.185 | 0.74 |
| Hepatitis B or C | Liver cancer | 0.75 | 1 | 0.654 | 0.768 | 3.154 | 0.52 |
| | | | 2 | 0.65 | 0.77 | 4.674 | 0.785 |
| | | | 3 | 0.65 | 0.77 | 4.674 | 0.785 |
| | | | 4 | 0.645 | 0.772 | 6.232 | 1.065 |
| | | | 5 | 0.641 | 0.775 | 7.866 | 1.37 |
| | | | 6 | 0.641 | 0.775 | 7.866 | 1.37 |
| | | | 7 | 0.641 | 0.775 | 7.866 | 1.37 |
| | | | 8 | 0.636 | 0.778 | 9.936 | 1.772 |
| | | | 9 | 0.636 | 0.778 | 9.936 | 1.772 |
| | | | 10 | 0.636 | 0.778 | 9.936 | 1.772 |
| | HCC | 0.82 | 1 | 0.702 | 0.771 | 1.777 | 0.227 |
| | | | 2 | 0.697 | 0.774 | 3.47 | 0.455 |
| | | | 3 | 0.697 | 0.774 | 3.47 | 0.455 |
| | | | 4 | 0.692 | 0.776 | 5.176 | 0.696 |
| | | | 5 | 0.686 | 0.779 | 6.967 | 0.961 |
| | | | 6 | 0.686 | 0.779 | 6.967 | 0.961 |
| | | | 7 | 0.686 | 0.779 | 6.967 | 0.961 |
| | | | 8 | 0.679 | 0.783 | 9.292 | 1.326 |

*First incident liver disease diagnosis was not cirrhosis, hepatitis fibrosis or sclerosis, but participants may have developed these conditions later

Analysis restricted to liver-related conditions with at least 5 HCC cases. Associations were estimated using Cox proportional hazard models with time as the underlying variable. Follow-up time was calculated from baseline blood collection for participants with prevalent disease or from age at first diagnosis if the condition occurred after study baseline, based on hospital admission records.

Cut-points derived using Youden's Index (maximum sensitivity + specificity – 1) at 3 years of follow-up
High risk(t) = Cumulative risk for participants with a protein risk score ≥ cutpoint (PPV(t))
Low risk(t) = Cumulative risk for participants with a protein risk score < cutpoint (cNPV(t))
Sensitivity(t)= Proportion of participants ≥ cutpoint who developed the outcome, relative to the mean risk in the population
Specificity(t)= Proportion of participants < cutpoint who did not develop the outcome, relative to the mean risk in the population.

Abbreviations: CI=confidence interval, HCC=hepatocellular carcinoma
